# Understanding Thromboembolus Transport Patterns In The Brain For Stroke In The Presence Of Carotid Artery Stenosis

**DOI:** 10.1101/2024.09.29.24314524

**Authors:** Ricardo Roopnarinesingh, Neel D. Jani, Michelle Leppert, Debanjan Mukherjee

## Abstract

Deciphering the source of an embolism is a common challenge encountered in stroke treatment. Carotid stenosis is a key source of embolic strokes. Carotid interventions can be indicated when a patient has greater than 50% stenosis in the carotid ipsilateral to the cerebral infarction, which is designated as the symptomatic carotid. However, there are often significant number of cases where carotid emboli travel contralaterally leading to ambiguity regarding which carotid is symptomatic. We use a patient-specific computational embolus-hemodynamics interaction model developed in prior works to conduct an in silico experiment spanning 30 heart-to-brain arterial models with differing combinations of bilateral severe and mild stenosis degrees. We used these models to study source-to-destination transport of thromboemboli released from left/right carotid disease sites, and cardiogenic sources. Across all cases considered, thromboemboli from left and right carotid sources showed non-zero contralateral transport. We also found that cardiogenic thromboemboli do not have an altered hemisphere distribution or distinct transport preference dependent on stenosis degree, thus potentially making the underlying etiology more cryptic. In patients with carotid stenosis or chronic occlusion ipsilateral to the area affected by stroke, we have demonstrated that the presence of contralateral stenosis can cause emboli that travel across the Circle of Willis (CoW) which can potentially lead to ambiguity when deciding which carotid is truly symptomatic.

## 1 Introduction

Embolic stroke accounts for a majority of all ischemic stroke cases, with some studies categorizing up to 30% of ischemic stroke cases as Embolic Stroke of Undetermined Source (*ESUS*) [1–3]. Currently, the ability to identify and disambiguate an etiology of a stroke is limited for ESUS cases, which can restrict treatment efficacy and lead to recurrent events [2]. Multiple co-existing potential sources is a common feature in ESUS cases that can make disambiguation of embolism source or etiology inherently difficult in ESUS cases. For instance, a common ESUS scenario may involve a patient experiencing both atrial fibrillation and carotid atherosclerosis ipsilateral to the occlusion location [3]. The carotid arteries cause 15%-20% of all ischemic strokes due to build up of atherosclerotic plaques that can generate thromboemboli that move into the cerebral arteries [4]. These buildups vary in severity based upon the degree of stenosis and laterality of the stroke which informs potential interventions [5–8]. Carotid interventions aimed at revascularization, either through carotid endarterectomy (*CEA*) or carotid artery stenting (*CAS*) are performed when there is stenosis to restore perfusion and reduce future stroke risk [9]. Specifically, carotid intervention is indicated when a patient has a North American Symptomatic Carotid Endarterectomy Trial (*NASCET*) severity greater than 70% in the carotid ipsilateral to the stroke along with transient ischemic attacks [7]. Carotid interventions are considered if the patient has a severity between 50% and 69% along with symptoms, but are not required as the benefits may be limited [7, 9]. Atherosclerotic plaque can become vulnerable to embolization without easily decipherable markers, and these vulnerable plaques can subsequently lead directly to a cerebral occlusion [10, 11]. Although stenosis severity may be non-trivial to acquire, it may not always be possible to tell where an embolus originated from due to the complexities of physiological arterial hemodynamics, and the complex embolus-hemodynamics interaction mechanics. It is commonly believed that emboli originating from a carotid artery will travel into the Circle of Willis (*CoW*) and occlude a vessel within the ipsilateral hemisphere, but there have been known cases where the occlusion occurs in the contralateral hemisphere [12–14]. Although contralateral events are less frequent, these cases can contribute to the ambiguity in deciphering stroke etiology due to the non-intuitive pathways of occlusion. With both carotids being suspect to combinations of vulnerable severe and mild levels of plaque buildup, contralateral events could lead to patients who receive treatment for the ipsilateral non-symptomatic vessel thereby deriving less benefit. These contralateral events can cause the chance of missing secondary prevention opportunities or possible benefits from intervening on the contralateral carotid if that is the truly symptomatic vessel. How contralateral carotid stenosis affects stroke risk is currently relatively unexplored, whereas this investigation can further illuminate mechanisms and risks of non-intuitive stroke in cases of bilateral stenosis [15, 16]. We additionally considered cases consisting of Contralateral Carotid Occlusions (*CCO*). Cases with CCO offer an interesting circumstance wherein each diseased carotid will be treated differently depending on which side is actively causing embolization. Understanding the probabilities of non-intuitive embolus source-to-destination movement for varying carotid stenosis cases can help to provide a more cohesive risk stratification across bilateral stenosis degrees. There is currently a lack of understanding in how patients with carotid disease may experience contralateral embolic events or non-intuitive embolic transport in various levels of carotid stenosis. Here, we present an *in silico* study analyzing how different combinations of varying carotid stenosis may present non-intuitive occlusion events and how the altered hemodynamics with the introduction of a stenosis can affect embolus transport.

## 2 Methods

### 2.1 Image-Based Modeling of Vascular Anatomy

A patient-specific vasculature model was created spanning arterial pathway from the aortic arch to the CoW using the open-source software tool Simvascular [17, 18]. The model was generated from a contrast enhanced computed tomography (*CT*) image, available from a set of CT images as part of the Institutional Review Board (*IRB*) approved Screening Technology and Outcome Project in Stroke (*STOP-Stroke*) database [19]. Since we used de-identified version of the CT image for secondary retrospective computational analysis, for this study no additional IRB approval was needed. The specific patient chosen for this study was based on the criteria that the patient exhibited a complete CoW anastomosis with healthy (*non-stenotic*) carotid arteries (*see also our prior works* [18, 20]). Arteries from the aortic inlet to the cerebral arteries up until the M1, A1, and P1 segments of the MCA, ACA, and PCA, were segmented using planar 2D segmentation technique built into SimVascular. These segmentations were then lofted to generate the 3D solid model of the complete arterial pathway. Arterial stenosis was virtually modeled at specific carotid artery locations, using these healthy patient vascular segmentations, and manually decreasing the segmentation sizes of the carotid vessels to the corresponding NASCET severity. The stenosis degrees were categorized into three mild NASCET scores of [10%, 25%, 40%], and three moderate/severe NASCET scores of [50%, 70%, 85%]. These severity ranks were paired such that one carotid would have a mild stenosis and the other would have a moderate/severe stenosis for both the left and right carotid arteries’ permutations. Additionally, by virtually eliminating these segmentations locally at the carotid site, we modeled 100% occluded carotid across all 6 severities for both carotids to mimic cases of CCO. Length of stenosis is defined as the span of artery that has a disturbed cross-sectional area due to the stenosis. For our modeling pipeline, the length of stenosis that spanned across the carotid artery varied proportionally with the severity of the stenosis. A more severe stenosis corresponds to a larger length. The equation of the stenosis length is defined in the Supplementary Material in greater detail. For all models in this study, we keep the stenosis geometry fixed. This is because, once thrombotic phenomena are initiated, the resulting clot may embolize without significant immediate arterial wall remodeling and modification in plaque geometry, especially within the short embolus simulation timescale as outlined in subsequent sections of our methodology. This procedure, therefore, led to 30 different models each with a unique pair of mild and moderate/severe stenosed carotids. The resulting models for the stenosis vascular anatomy for differing severities can be seen in Figure 1.

**Figure 1:**
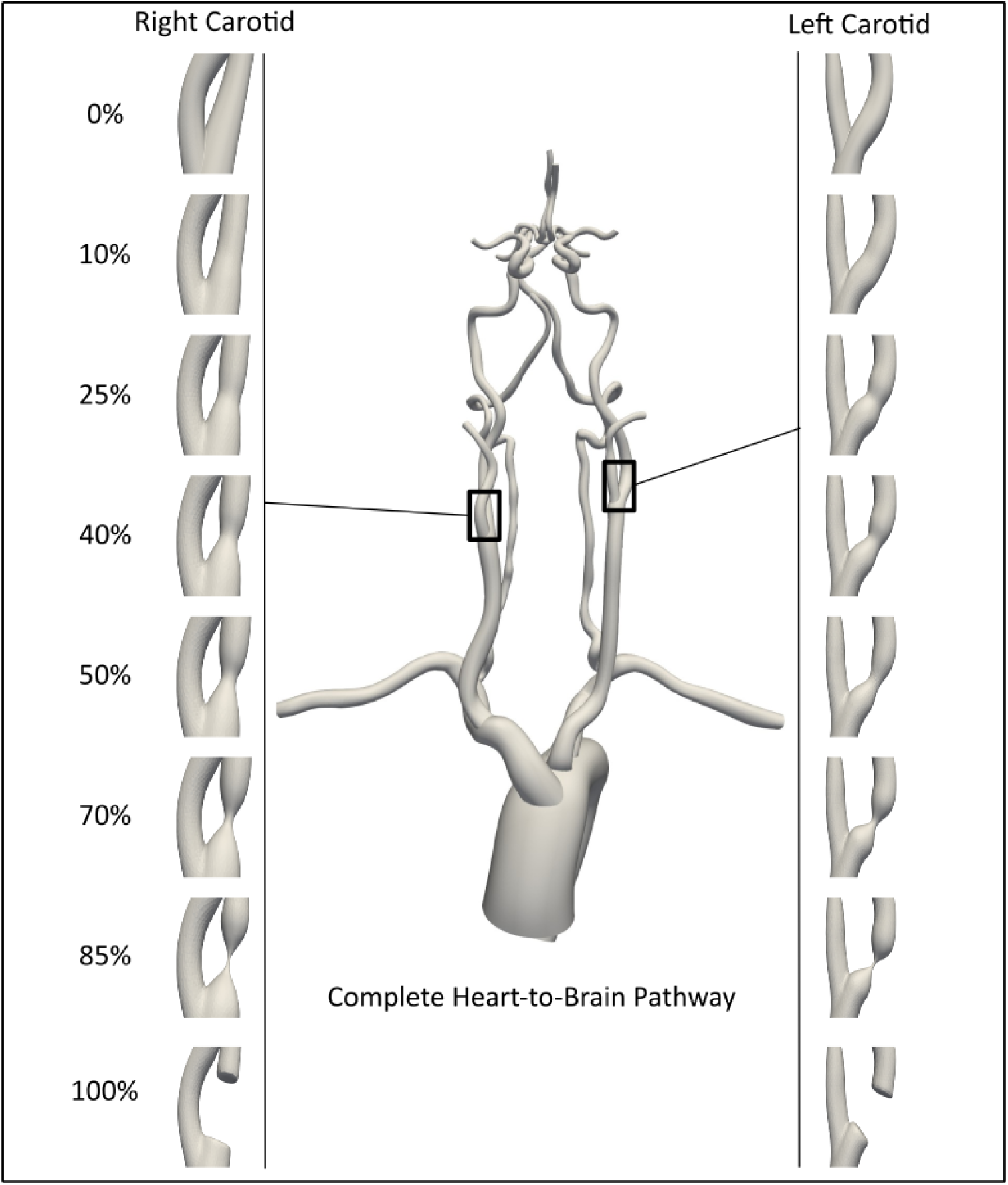
An illustration of the varying stenosis models for the right and left carotid. All stenosis degrees were imposed onto a single set of patient imaging data with pairings of non-severe and severe carotid stenosis degrees applied to each carotid. The 100% severity, or contralateral carotid occlusion cases, were paired with each of the non-severe and severe stenosis degrees.

### 2.2 Hemodynamics Simulation

Hemodynamics across the arterial network model was simulated using a stabilized finite element method for incompressible Navier-Stokes equations implemented in SimVascular [17]. Each model was discretized into a mesh comprising 9-10 million linear tetrahedral elements. Three levels of mesh sizing were specified. First, guided by mesh refinement analysis described in prior works [21], an average element size for the entire network was specified across all models to be 0.65 mm. Second, local refinement operation was conducted for vessels in the CoW, with an average element size of 0.25 mm due to a smaller cross-sectional area relative to other vessels in the heart-brain arterial model. Third, stenosed regions of the carotid arteries underwent additional element refinement due to the smaller cross-sectional area. This local refinement was guided by setting the element edge length to 1/10th of the smallest stenosis diameter, ensuring at least 10 or more elements within all cross-sectional slices of the stenosed region. Blood was assumed to be an incompressible Newtonian fluid with average viscosity of 4.0 cP, and bulk density of 1.06 g/cc. Mathematical details of the underlying equations for solving flow velocity and pressure using stabilized finite elements are provided in the Supplementary Material. A physiological pulsatile flow profile obtained from magnetic resonance imaging measurements with a cardiac cycle of 0.83 seconds was applied at the aorta inlet [22]. This inlet flow was set to a consistent cardiac output (CO) of 79 mL/s across all models. Resistance boundary conditions were assigned to each outlet of the model to account for the influence of the downstream vascular beds at the arterial outlets. For this purpose, Total Arterial Resistance (TAR) was estimated using a Mean Arterial Pressure (MAP) of 93.33 mmHg, which assumes that the patient exhibits an average systolic and diastolic blood pressure of 120 and 80 mmHg respectively. A proportion of the TAR was assigned to each vessel outlet based on target flow divisions as outlined in prior work [18, 20, 23]. For target flow division, 65% of the total CO was assumed to exit the descending aorta [24], flow rates exiting the six cerebral artery outlets were propotionately assigned based on measured MR data reported in [25], and the remainder volumetric flow was set to exit the external carotid and subclavian arteries proportional to their cross-sectional areas [26]. These resistance values were tuned to the targeted flow values through multiple steady-state flow simulations as outlined in [23, 27]. Once obtained, the CO, TAR, MAP, and outlet resistances were kept the same across all models to enable controlled simulations of embolus transport with varying stenosis severities only. Hemodynamics was simulated for three cardiac cycles for each model, and computed velocity and pressure fields for the third cycle were used to conduct embolus transport simulations. Additional details of these hemodynamics simulation steps are outlined in our previous works [18, 20, 23], and have not been reproduced here for conciseness of presentation.

### 2.3 Embolus Transport Simulation

Embolus transport was modeled using a custom modified version of the Maxey-Riley equation [28] to represent embolus-hemodynamics interactions, established extensively in our prior works [27]. This model estimates individual embolus velocity and position as it traverses through blood flow based on embolus properties and hemodynamic forcing. To closely simulate arterial embolus transport, we modified the classical form of Maxey-Riley equations to account for: (a) lift forces produced by shear gradients near artery walls; (b) collisions between particles and the artery wall; and (c) elastohydrodynamic lubrication effects between embolus and artery wall. These forces were in addition to the forces of drag, added mass effects, and forces from the undisturbed background flow. We note that no additional cross-stream dispersion forces due to margination effects were incorporated here, owing to significant size and inertia differences between blood cellular components and the embolus, as well as evidence of margination effects being weak in large vessels, especially considering substantial radial flow components originating from vessel curvature and tortuosity [29, 30]. Mathematical details of the underlying equations are provided in the Supplementary Material. The emboli were assumed to be spherical particles with negligible deformations (*i.e. their radius remains constant*), with a one-way coupled fluid-particle interaction model (*that is, while fluid forces drive the emboli, the emboli in turn do not significantly alter the flow around them*). Individual embolus size was set to be 500*µ*m, which leads to a momentum response time of 0.004 sec, and a Stokes number (*compared to cardiac cycle duration*) of 0.005. This size was chosen to focus on small emboli whose transport can be more ambiguous to infer from imaging; while remaining within the bounds of the mathematical approximations of one-way coupled embolus hemodynamics interactions, which break down as embolus size approaches vessel diameter [31]. Individual emboli were assigned mechanical properties of that of thromboemboli as obtained from literature [27], and this material behavior was accounted for in the model when resolving embolus collisions with vessel wall. Specifically, a viscoelastic, velocity dependent restitution coefficient accounting for corrections due to elasto-hydrodynamic lubrication effects was used to resolve the embolus interactions with the vessel wall [27]. We note that individual embolus-to-embolus collisions were not modeled here as each simulation comprises individual embolus trajectory estimation through a Monte Carlo sampling-based approach detailed in the next section. The simulated flow field for the third cardiac cycle (see Section 2.2) was stitched together across 10 cardiac cycles to generate a representative hemodynamic field with no cycle-to-cycle variation. 10 cycles was chosen, based on observations from prior numerical experiments, to allow enough time to distribute all emboli to an outlet, leaving less than 10% of emboli samples recirculating in the vessel network (*unresolved emboli*). For instances where more than 10% unresolved emboli were obtained after 10 cardiac cycles, the trajectories were evolved for another 10 cycles to ensure that the number of unresolved emboli samples is always less than 10%. Individual thromboembolus trajectories were calculated by numerical integration of the custom embolus dynamics equation using a Euler integration scheme with a numerical time step of 0.05 ms. Supplementary Animation V2 presents an illustrative example of source-to-destination embolus transport dynamics computed for one of the stenosis models.

### 2.4 Design of In Silico Experiments

We utilized the simulation methodology introduced in Sections 2.1-2.3 to structure a parametric set of *in silico* experiments to investigate embolus transport in the presence of varying carotid stenosis severities. The outline of this experiment design is shown in Figure 2. To parameterize the stenosis severity, a total of 18 vascular models were created as a combination of one mild stenosis (*10%, 25%, and 40% NASCET severity*) and one moderate/severe stenosis (*50%, 70%, and 85% NASCET severity*) for each pair of left and right carotids respectively (*see also Figure 1*). An additional 12 models were designed with a fully occluded carotid artery, mimicking CCO cases, paired with one of each of the mild and severe stenosis degrees. For each of these 30 models, approximately 5,500 thromboemboli samples were released along the carotid artery walls, mimicking embolization at the stenosis site. For the 12 CCO models, thromboemboli samples were released at the partially stenosed carotid site; while for the 18 non-CCO models, thromboemboli samples were released from both carotid sites bilaterally. For all carotid surface releases of emboli, the individual particles were offset by a distance equal to the embolus radius, to avoid numerical artifacts due to particles remaining stuck at the walls. An illustration of this release configuration is presented in Supplementary Material, in Figure S1. Furthermore, for all 30 cases, another ≈ 5,500 samples of cardiogenic thromboemboli were released at the aortic root inlet, to further illustrate how different combinations of stenosis severities may affect the cardiogenic emboli pathway towards the cerebral arteries. This design led to a set of 78 total embolus transport experiments, leading to combined distribution data spanning approximately 429,000 thromboemboli. (Supplementary section S5 further shows surface release configurations for the cardioembolic and carotid sources).

**Figure 2:**
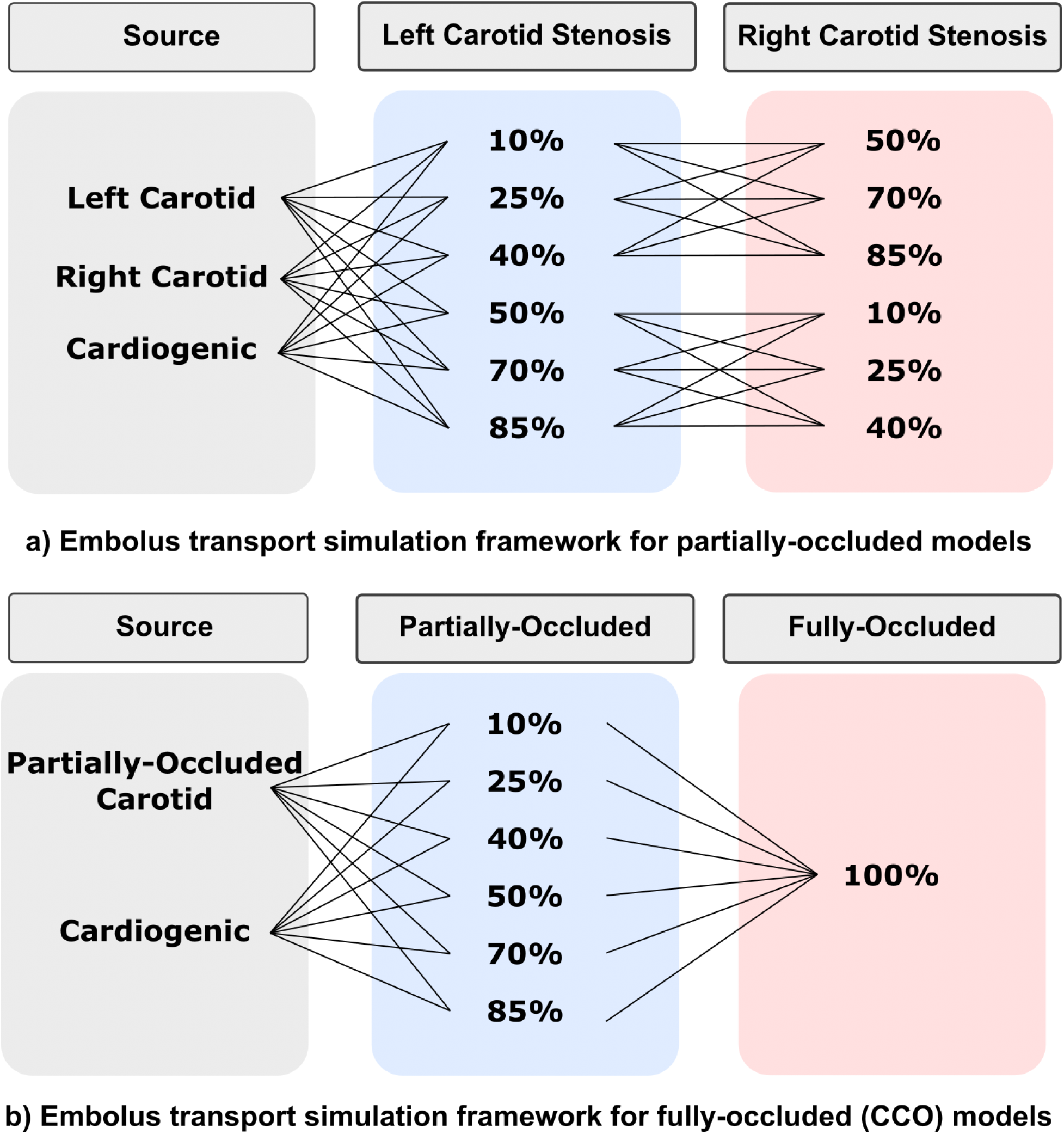
Design of the in silico experiments for embolus transport. Two sets of models were created for these simulations. a) Partially-occluded models were built using pairing of non-severe and severe stenosis degrees across both the left and right carotid. Emboli were released from the both carotids and the face of the aortic root for cardiogenic cases. This totaled to 18 unique models with differing pairing of severe and non-severe stenosis degrees with embolus releases from these three sources. b) 12 models were created in which one of the carotid arteries were fully-occluded (100% NASCET degree) and the other carotid was partially occluded. These pairings were completed to where there was one fully-occluded carotid and the other carotid was partially-occluded using each degree of severe and non-severe NASCET rankings. Emboli were released from the partially occluded carotid and the face of the aortic root.

### 2.5 Data Analysis

For each experiment, the trajectory of each individual embolus sample was tracked from release source to a destination vessel, and number fractions of samples distributed to the 6 cerebral arteries was computed and analyzed for this study. The number of emboli reaching vessel outlets were counted at the left and right middle cerebral artery (MCA), left and right anterior cerebral artery (ACA), left and right posterior artery (PCA), as well as the left and right subclavians and external carotids, and the descending aorta. Additionally, hemispheric distributions were calculated by separating the left and right embolus counts within the ACAs, MCAs, and PCAs to analyze contralateral or trans-hemispheric distribution occurrences. In addition, for each of the vascular model, the outlet flow rates to each of the outlets listed above were also computed. The resulting flow and embolus distribution data was compared to assess key trends in embolus source-to-destination mapping. These comparisons were evaluated using statistical tests, primarily non-parametric tests such as Mann-Whitney U and Wilcoxon. The non-parametric tests were chosen since they do not require the underlying sample data to be normally distributed, which we assessed using the Shapiro-Wilk test for normal distribution. Specifically, a non-parametric one-sample Wilcoxon t-test is utilized to establish whether significant levels of non-intuitive contralateral or trans-hemispheric embolus transport occurs per carotid embolus release; and to compare flow rate differences within the communication arteries in the Circle of Willis, between the stenosis models and a baseline model without stenosis. Additionally, a Mann-Whitney U test was used to compare the likelihood of contralateral movement in mild/moderate stenosis degree releases to those in severe stenosis degree releases; as well as to compare the likelihood of contralateral movement between fully-occluded (CCO) models and partially occluded models.

## 3 Results

### 3.1 Trends in Contralateral Distribution of Carotid Thromboemboli

Thromboembolus distribution from our simulations was computed as the percentage of emboli that exit each of the six outlets of the CoW. Figure 3 presents the proportion of emboli that were able to cross into the opposite hemisphere relative to the carotid release site for each pair of stenosis severities, illustrated for the 18 bilateral stenosis, non-CCO cases. Each model is labeled by their combination of left-right stenosis severity respectively (*i.e. 10L50R corresponds to the model with a 10% left carotid stenosis and a 50% right carotid stenosis*). These embolus distribution results therefore inform a probability of contralateral embolic events occurring with respect to stenosis severities across the right and left carotid arteries. We observed that, all simulations produced a non-zero contralateral distribution, meaning that all combinations of severe and mild stenosis showed the potential for contralateral occlusions. The largest extent of contralateral movement were found in the models with the most severe (*that is, 85%*) stenosed carotid, where contralateral movement was highest from the mild carotid release sites. These models had a likelihood of contralateral distribution between 6% and 10%, which was notably higher than the other releases that ranged primarily between 0.5% and 4%. Statistically, the null hypothesis that contralateral movement does not occur from emboli released at both the left and right carotids (*that is the proportion of contralaterally traveling emboli = 0*) was rejected with p-values both *<* 0.01 (*based on a one-sample non-parametric Wilcoxon rank test*). We also observe that extent of contralateral movement of thromboemboli is generally greater for emboli released in right carotid and traveling towards the left hemisphere for severely stenosed left carotids, indicating an inherent right vs left asymmetry for the cases considered here. Additionally, figures S2 and S3 in supplementary section S5 present complete outlet distributions that illustrate arterial regional preferences for each model. Generally, these simulation data revealed contralateral trans-hemispheric thromboembolus transport and distribution occurring at the CoW; with primary modality of embolis moving from the mild stenosis site towards the hemisphere ipsilateral to the severe stenosed site, and with the likelihood of contralateral movement generally increasing as the moderate/severe carotid stenosis increases in degree. Animation V3 in the supplementary section presents contralateral movement occurring in the 40% right carotid occlusion and 85% left carotid occlusion model. Additionally, animation V4 visually depicts how contralateral movement is dependent on the bilateral degree of stenosis in which the propensity for contralteral movement differs.

**Figure 3:**
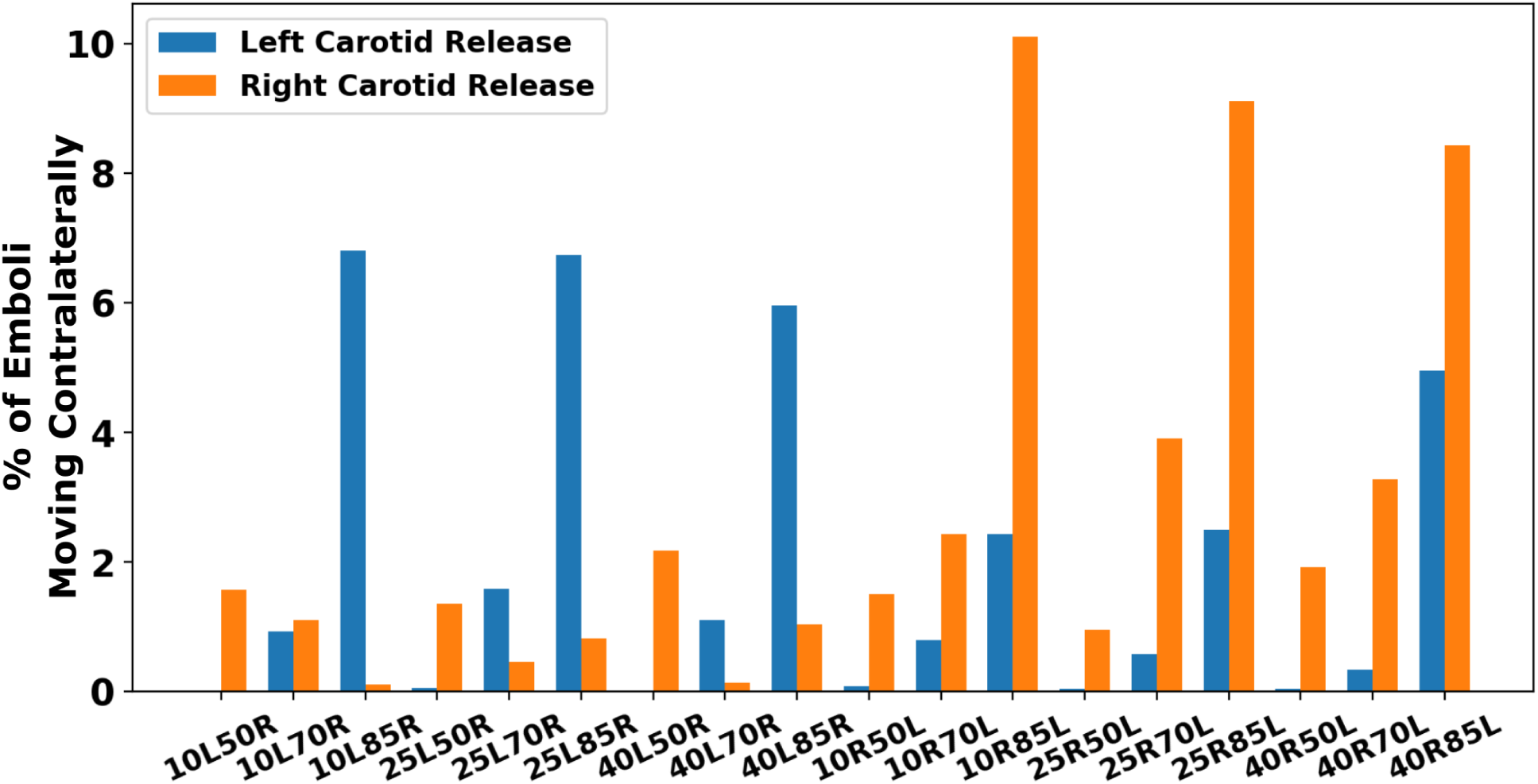
The proportion of emboli that completed contralateral movement for each pairing of stenosis degrees. The x-axis labels each model with their respective left and right stenosis degree (i.e. 10L50R indicates the model with a 10% left carotid stenosis and a 50% right carotid stenosis).

### 3.2 Trends in Cardiogenic Thromboembolus Distribution

In Figure 4, we present the cardiogenic thrombembolus distribution to each cerebral hemisphere relative to each severity combination, for the 18 bilateral stenosis, non-CCO cases. The individual models are labeled using the same scheme as in Figure 3. The cardiogenic distribution was acquired to illustrate how the thromboembolus trajectory and their source-to-destination propensities could get modified based on the severity of the stenosis. Thromboemboli traveling from the aortic root will be recruited into either the subclavian artery or the left/right common carotid artery and distribute into each hemisphere through the supplying cerebral arteries in the CoW. The resulting cardiogenic embolus distribution can help illuminate preferential pathways in the presence of varying stenosis degrees for both the right and left carotid arteries that can, in turn, reveal non-intuitive pathways which potentially contribute to ESUS diagnosis. Our experiments reveal a rather consistent distribution of emboli across all models averaging 55% and 45% for the left and right hemispheres respectively (*standard deviation of 4.78%, coefficient of variation for left* = 0.087*, for right* = 0.106). These observations indicate that stenosis severity may not potentially have a strong influence on distribution of cardiogenic thromboemboli across both hemispheres. This itself is counter-intuitive, as specifically for the high severity cases, it may seem expected that hemisphere ipsilateral to the high severity stenosis will not be a preferred cardiogenic embolus distribution. These observations therefore indicate the potential role of compensation from other vessels feeding the CoW, and the overall network anastomosis of CoW, by which cardiogenic thromboemboli are able to distribute into the hemisphere with a severely stenosed carotid artery. This is further explored next (Section 3.4) by comparing computed volumetric flow rates feeding into the cervical vessels that supply the CoW. Figure 5 illustrates the total embolus distribution from cardiogenic sources as they traverse the left and right carotid vessels, indicating clearly that cardiogenic thromboemboli have a marked propensity to travel along the right carotid pathways. This right side preference is commonly understood from an anatomical standpoint (*right cervical vessels branch out from the aortic arch first, followed by the left cervical vessels*), and has been observed across several prior studies [23, 32], thereby adding an indirect validation point for our simulated dataset. Additionally, supplementary Figure S4 provides total cardiogenic distributions to all outlet vessels in which no general regional preference is noted with slight variation for high stenosis degree.

**Figure 4:**
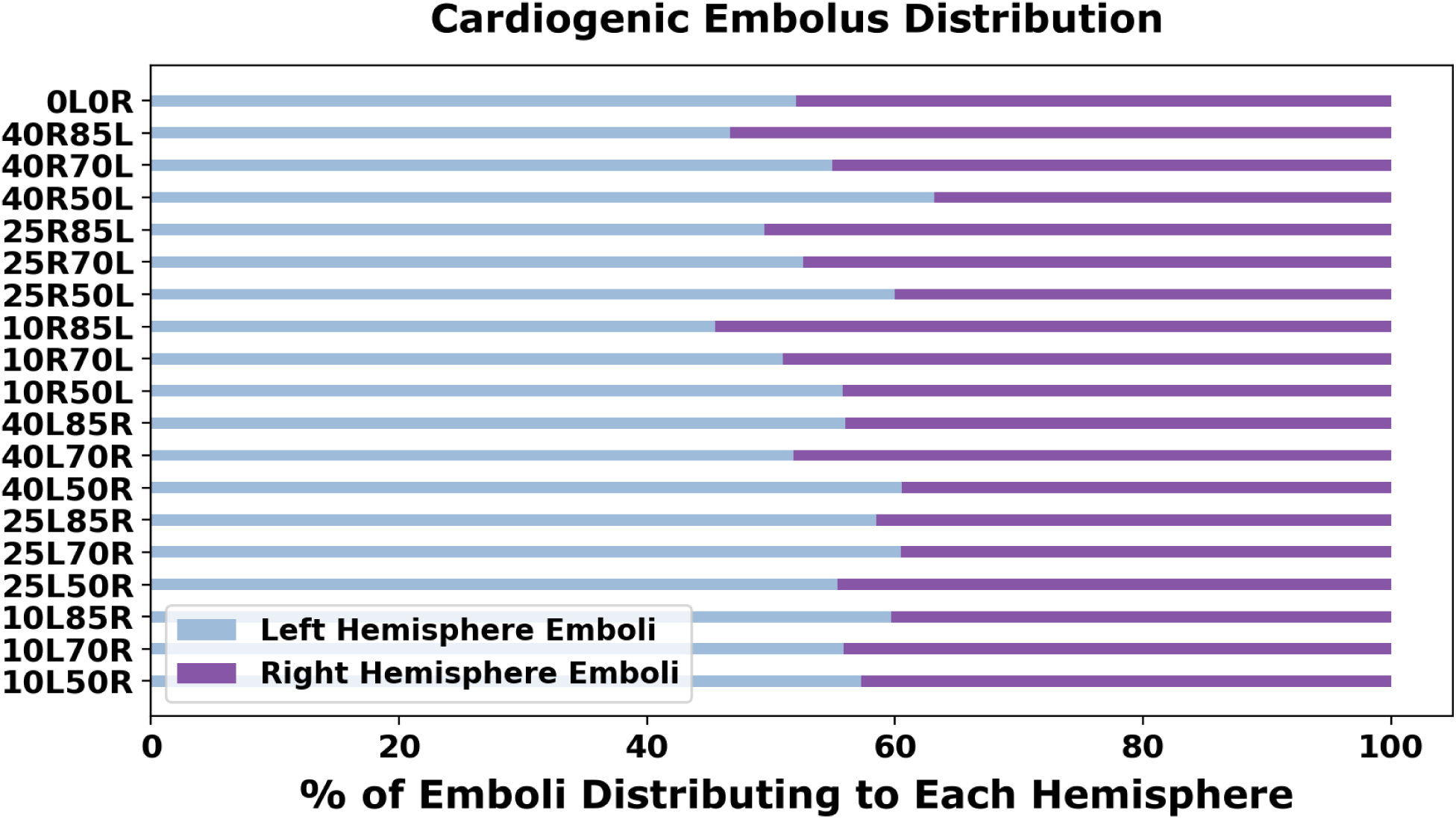
Percentage distribution of cardiogenic emboli to each CoW hemisphere. The y-axis labels each model with their respective left and right carotid stenosis degree (i.e. 10L50R indicates the model with a 10% left carotid stenosis and a 50% right carotid stenosis).

**Figure 5:**
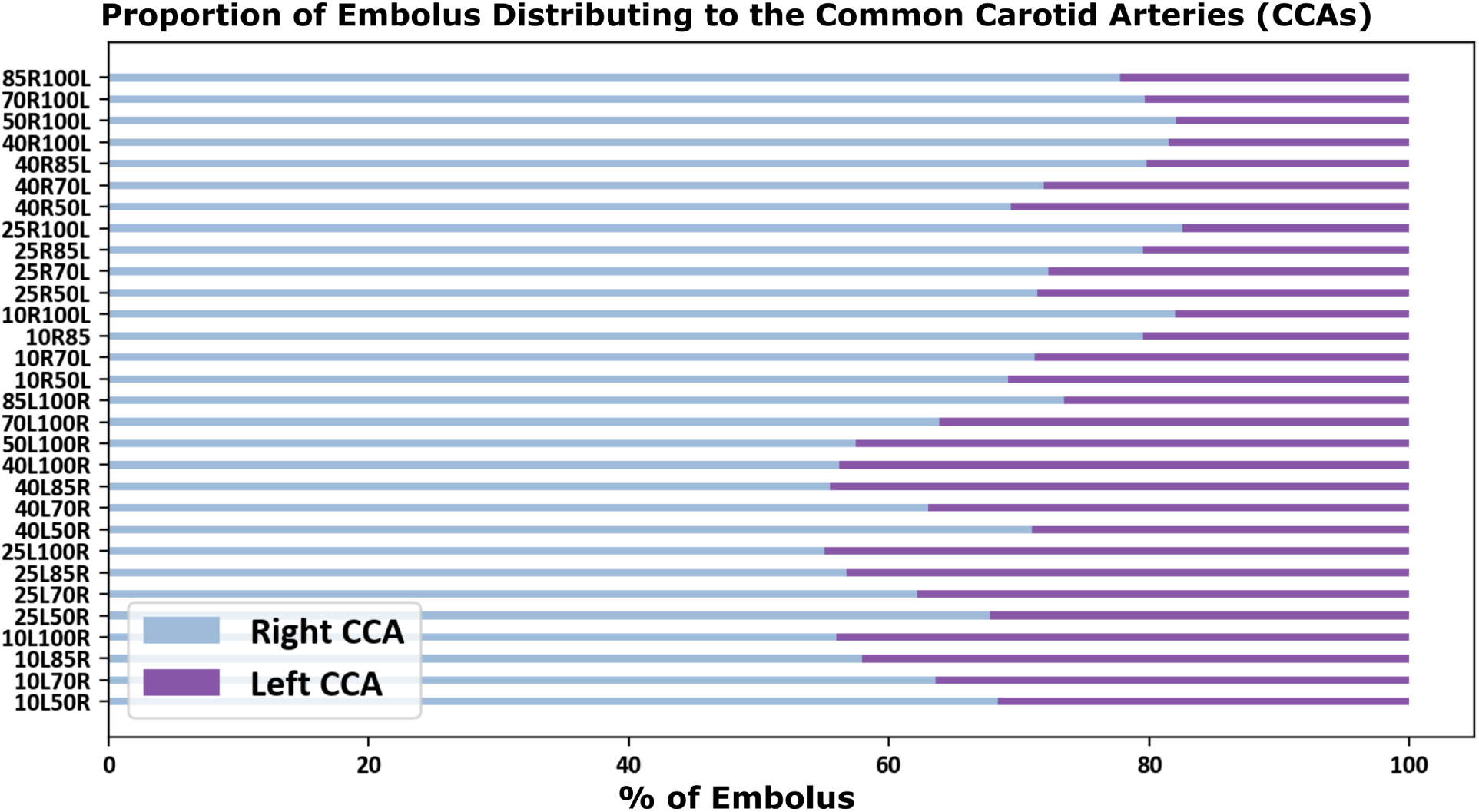
Distribution of cardiogenic embolus into the left and right common carotid arteries (CCAs). The y-axis denotes the bilateral stenosis degrees with the x-axis providing the percentage of embolus distributing into both of the CCAs. Embolus have a preference of traveling into the right CCA versus the left.

### 3.3 Thromboembolus Distribution in Contralateral Carotid Occlusion Cases

We further present thromboembolus transport and distribution in the 12 CCO models considered in our study. For this, we considered emboli releases from the carotid artery site that was not fully-occluded, and from aortic inlet (*that is, cardiogenic*). Figure 6 illustrates the proportion of carotid thromboemboli with contralateral trans-hemispheric distribution relative to the carotid release site for the 12 CCO models. The observations indicate a similar continuing trend compared to that observed in the results presented in Section 3.1, wherein increasing stenosis severity on one side permits increased contralateral movement for emboli released on the other side. The proportions indicated are for emboli reaching the hemisphere ipsilateral to the fully occluded carotid. The completely occluded cases allowed for a high proportion of emboli traveling contralaterally from the non-occluded carotid side with an average of 10.8% of contralateral emboli which is higher than all of the non-CCO simulation results presented earlier (*two-sample, Mann-Whitney U test between CCO cases and non-fully occluded cases: p-value <* 0.05). Furthermore, generally contralateral movement of carotid emboli in the simulated results are greater for 100% occluded left carotid compared to the right, indicating potential right vs left asymmetries as noted in Section 3.1. Furthermore, we also simulated cardiogenic thromboembolus and distribution for the 12 CCO cases, and the resulting distributions are presented in Figure 7. Cardiogenic thromboembolus distribution showed lesser variations in left vs right hemisphere distribution when compared against carotid emboli, especially for the fully occluded right carotid cases. The fully occluded left carotid models saw an average of 57.4% of emboli going to the right hemisphere and 42.6% going to the left (*standard deviation of 7.95%, coefficient of variation for right and left* = 0.14 *and* 0.19 *respectively*) while the fully occluded right carotid models experienced an average 52.1% distributing to the left hemisphere and 47.9% distributing to the right (*standard deviation of 3.04%, coefficient of variation for right and left* = 0.063 *and* 0.058 *respectively*). Thus, while it was favorable for emboli to distribute to the hemisphere fed by a non-occluded CCA, the variability in the split of left and right hemisphere thromboembolus distributions remained low regardless of the stenosis severity of the non-occluded carotid. These observations again indicate potential role of flow pathways where the non-occluded cervical vessels (*e.g. left carotids and basilar arteries, for fully occluded right carotids*) feeding the CoW can recruit emboli towards the brain, leading to non-intuitive cardiogenic embolus transport patterns. Animation V5 supplied in the supplementary section illustrates the differing transport mechanics of the embolus dependent on the degree of partially-occluded carotid. The depiction presents both release locations of the aortic root and the non-fully occluded carotid.

**Figure 6:**
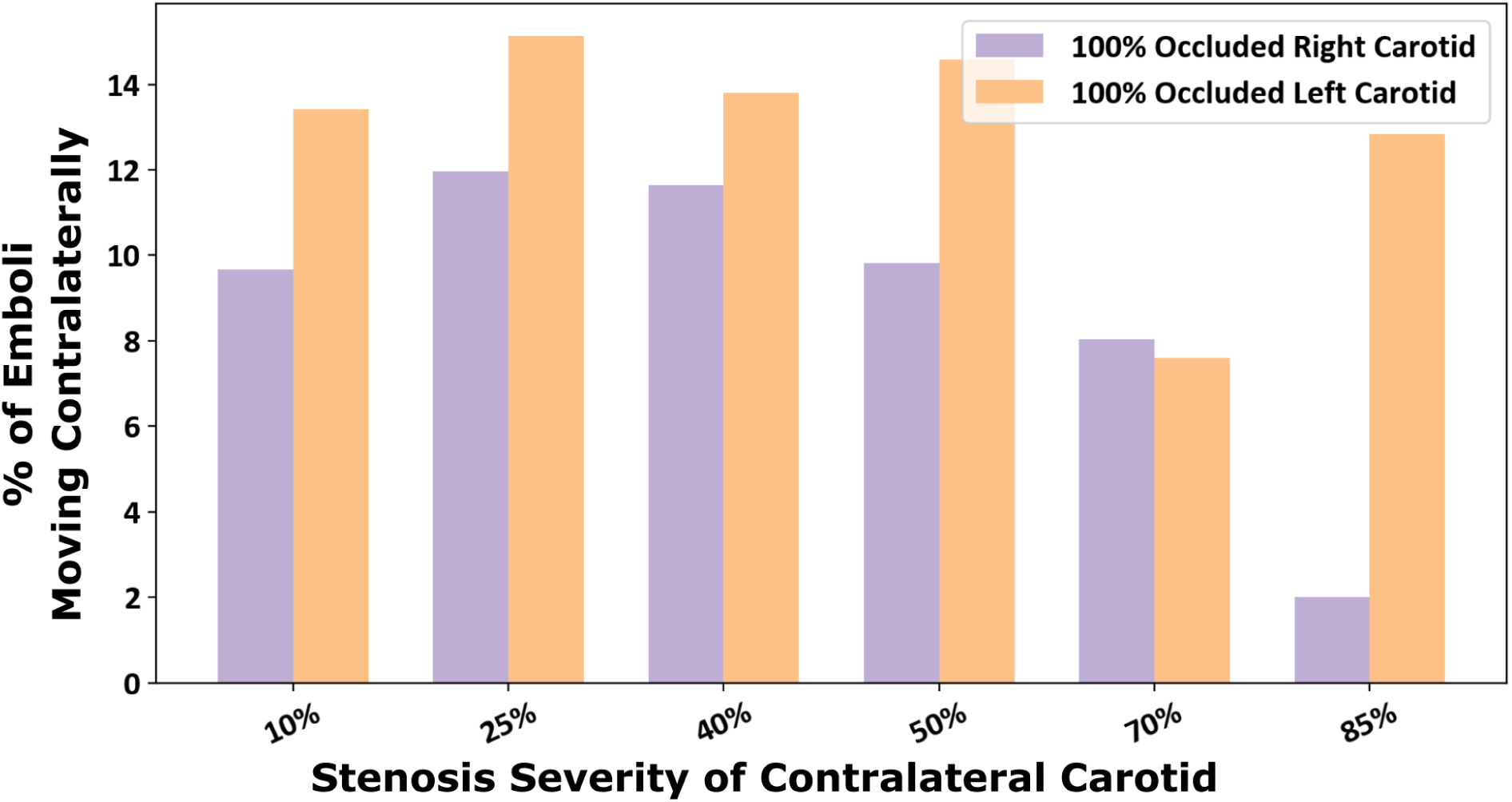
Proportion of emboli that distributed contralaterally within the models with a fully-occluded carotid artery (contralateral carotid occlusion or CCO cases). The x-axis indicated the stenosis severity of non-fully-occluded carotid with each bar designating a fully occluded left or right carotid.

**Figure 7:**
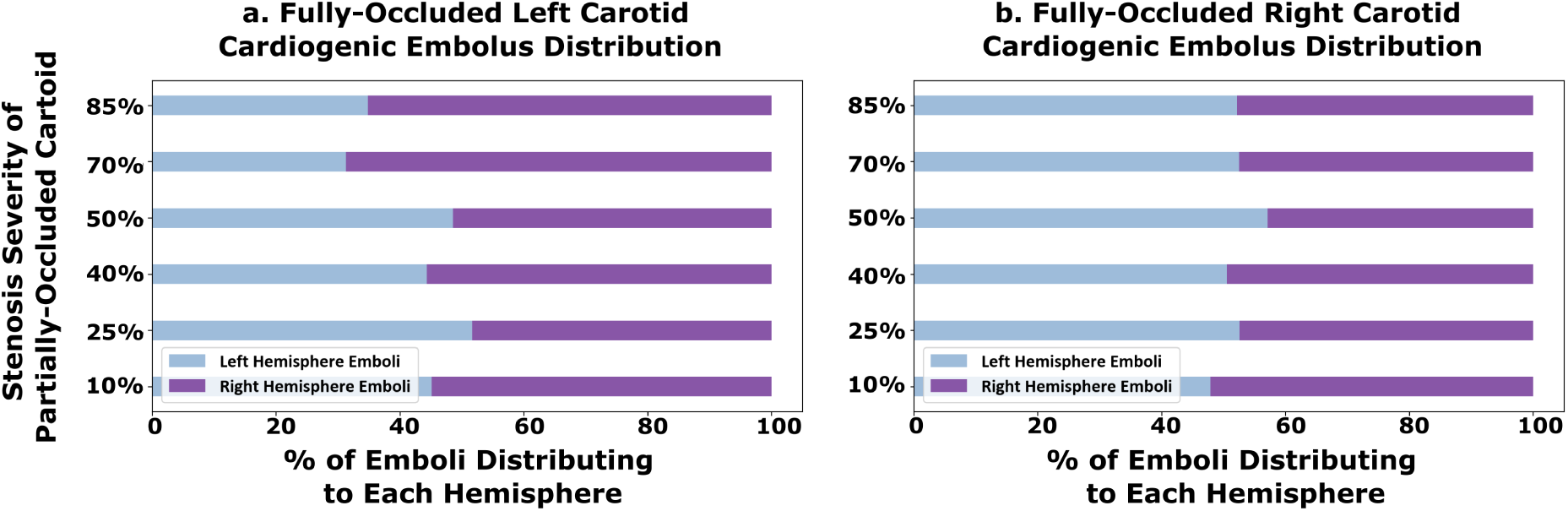
The proportion of cardiogenic emboli that travel to each cerebral hemisphere arteries of the models with a fully-occluded carotid (CCO cases). Panel a. (left) shows the hemispheric-embolus distribution for models with a full-occluded left carotid, and panel b. (right) shows the same for models with a fully-occluded right carotid. The y-axis indicates the stenosis severity of the contralateral carotid artery (which is not fully occluded).

### 3.4 Flow Distribution in Vessels Feeding the Circle of Willis

We further explore the network effect of the vessel anastomosis connecting the cervical vessels to the CoW, by analyzing estimated flow rates in the cerebrovascular network for all the 30 models. For all analyses presented here, flow rates were estimated by integrating the simulated blood flow velocity over a cross-sectional slice of the specific artery at each simulated time step. The total average flow rate was then computed by integrating the flow rates in time over one full cardiac cycle. Figure 8 illustrates the average volumetric flow rate through the cervical vessels feeding the Circle of Willis (*basilar artery, the left internal carotid artery, and the right internal carotid artery*), computed for the 18 non-CCO models with patent bilateral stenosis as proportions of total cerebral blood flow (*tCBF*). Likewise, Figure 9 presents the estimated flow rates at the basilar, and left and right carotid arteries for the 12 CCO cases, computed as proportions of total cerebral blood flow (*tCBF*). The computed hemodynamics as illustrated in Figure 8 for the 0% left stenosis and 0% right stenosis (*i.e. 0L0R*) case, agree well with reported cohort averaged blood flow distributions obtained from magnetic resonance imaging (MRI) scans in prior work [33]. Specifically, in prior measurements, the reported RICA, LICA, and BA, flow rates are: 36 4%, 36 4%, and 29 10% of the tCBF [33]. Flow-rates obtained from our simulations show RICA, LICA, and BA flow rates of 39%, 37%, and 24% (*shown in Figure 8*) which agree well with the MRI measured flow distribution. Furthermore, Figure 10 show the final flow distributions across all outlet vessels of the cerebrovascular network. and we observe that the flow distributions to the cerebral outlets (*as well as others*) remains consistent regardless of stenosis severity within the carotid arteries. This corroborates with our specification of controlled or fixed outflow boundary conditions across all models, and enables some insights on potential variations in flow routing across the vascular network with carotid stenosis. In particular, despite consistent outflow rates, the flow rate variations with varying stenosis severity at the cervical vessels, indicate that owing to the network nature of the vascular anastomosis leading into the CoW, the three cervical vessels feeding the CoW can be engaged differently for varying stenosis severities to retain consistent volumetric flow into the brain. This provides one mechanism of non-intuitive distribution of emboli. Specifically, increasing right carotid flow for severely stenosed left carotids will lead more emboli from the right carotid site to reach the CoW, increasing the likelihood of their contralateral movement. This is despite there being a faster jet flow at the severe stenosis site that locally increases the speed of any emboli released at the severe stenosis carotid site. An animation of differing jet flow within the stenosis region dependent on the contralateral carotid occlusion is included in the supplementary section video V1 illustrating how the network hemodynamic effects result in bilateral flow changes. Additionally, flow compensations within the basilar artery and the mildly stenosed carotid, to preserve cerebral perfusion, can pose as an alternate pathway of cardiogenic thromboemboli into the hemisphere ipsilateral to the severely stenosed carotid. Together, these observations indicate flow distribution and recruitment as a mechanism of consistent hemisphere distribution for cardiogenic emboli regardless of stenosis severity.

**Figure 8:**
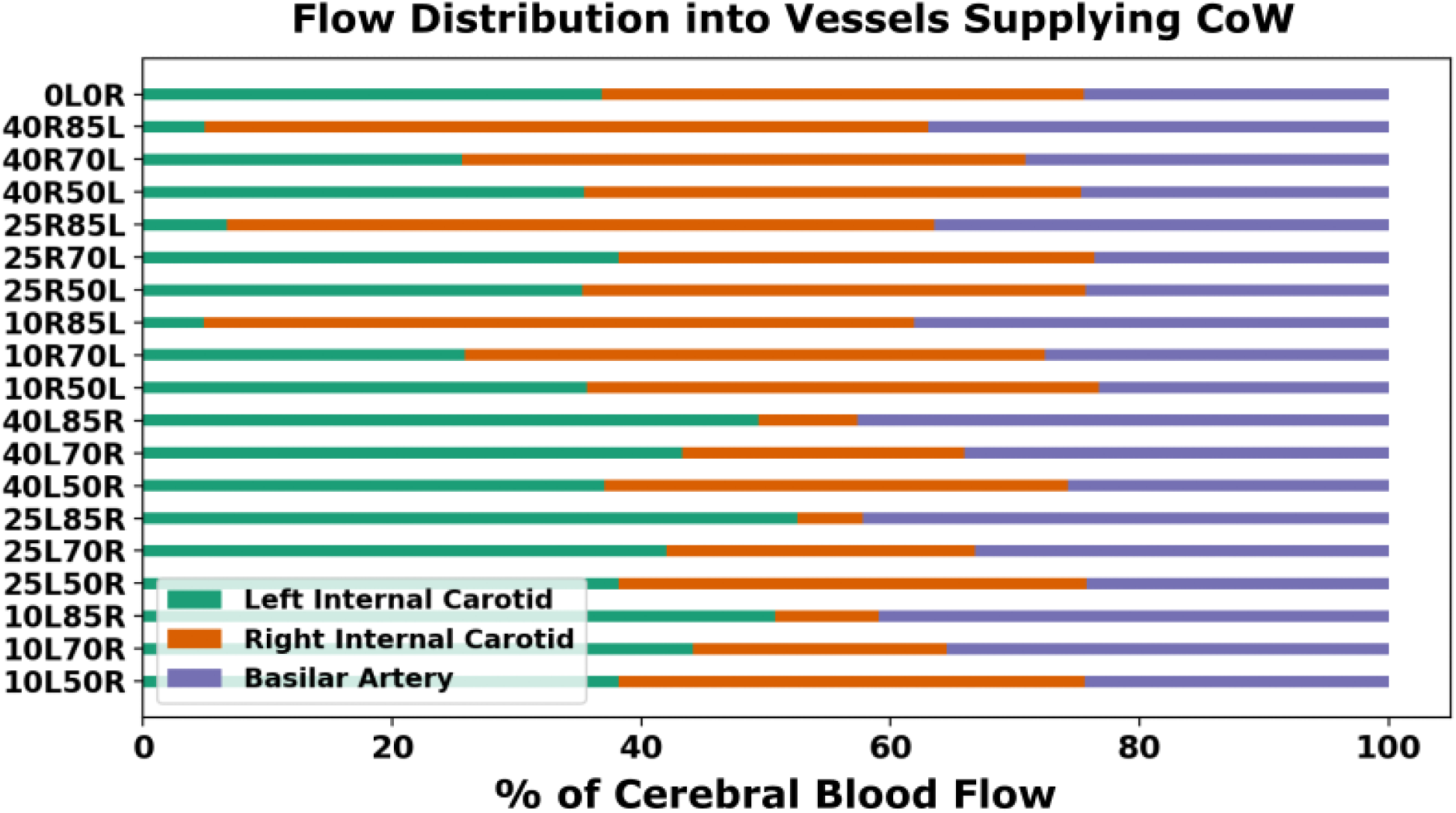
Percentage distribution of volumetric flow rate through each of the vessels feeding the Circle of Willis: the left internal carotid (LICA), right internal carotid (RICA), and basilar artery (BA). These distributions are illustrated across all combinations of severe and non-severe stenosis degrees where the basilar artery and less severely stenosed carotid artery compensate for flow as the carotid with severe stenosis increases in degree. Note that for the baseline no-stenosis model (0L0R), computed flow-rates into the LICA, RICA, and BA are: 39%, 37%, and 24% respectively, which agrees well with averaged MRI measurements reported in prior works [33].

**Figure 9:**
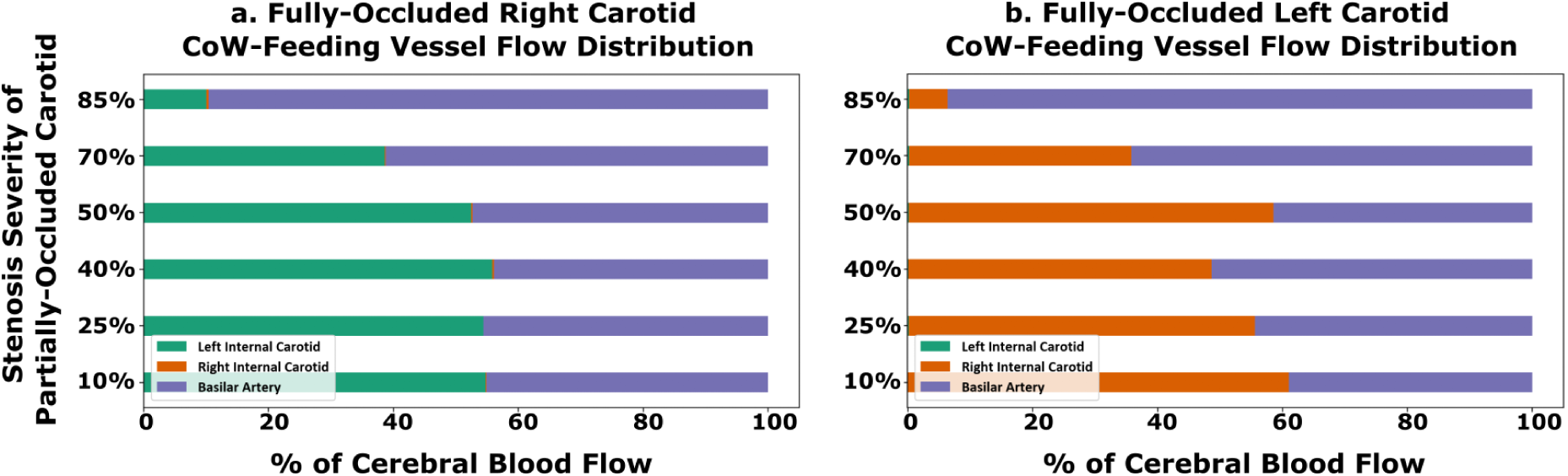
Percentage distribution of volumetric flow rate through each of the three vessels feeding the Circle of Willis for the cases with fully-occluded carotid artery. Panel a. shows the distribution in terms of total Cerebral Blood Flow (tCBF) for models with a fully-occluded right carotid and panel b. shows the same for models with a fully-occluded left carotid. The y-axis indicates the stenosis severity of the contralateral carotid artery (which is not fully occluded).

**Figure 10:**
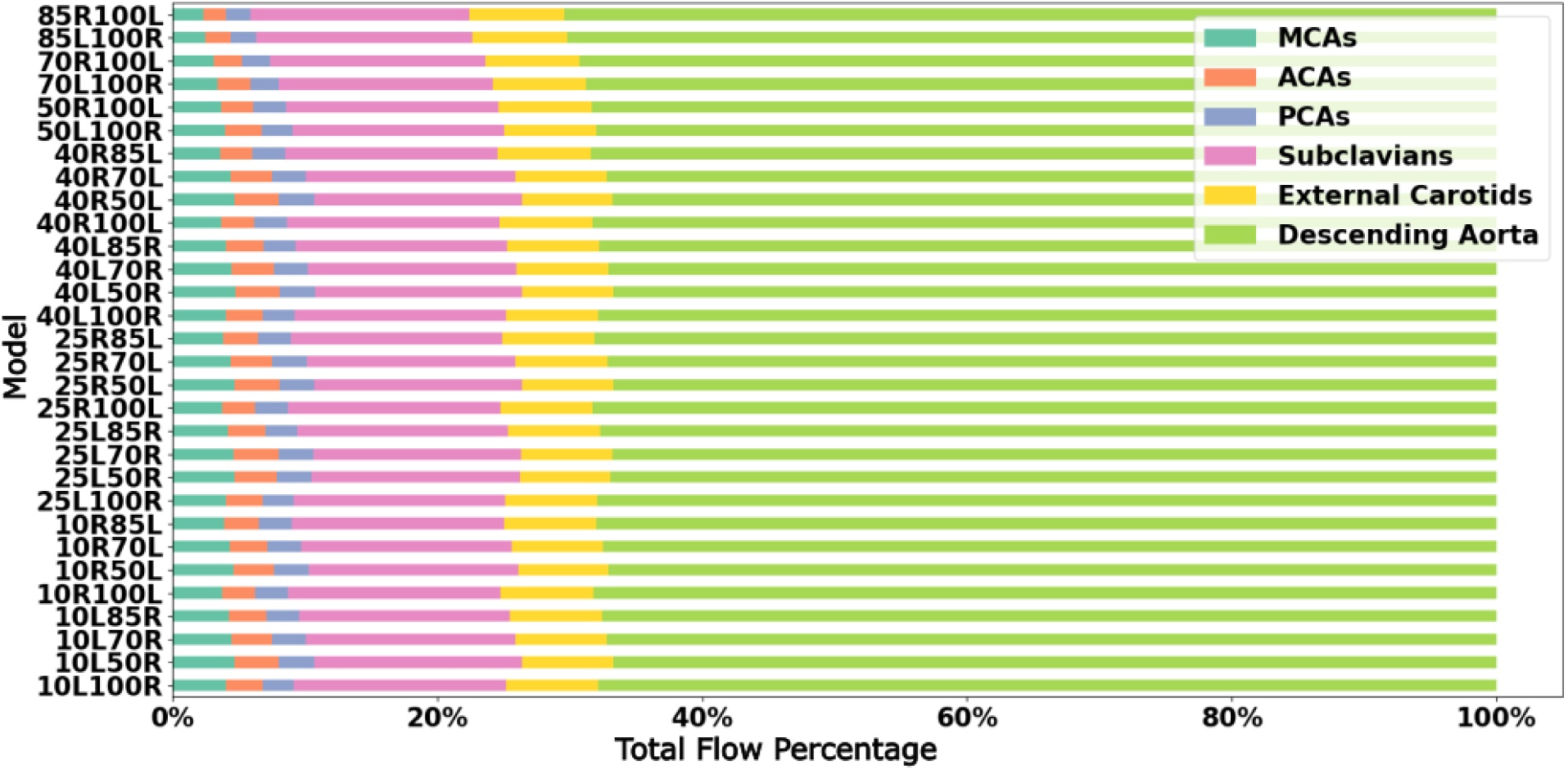
Average volumetric flow rates to the outlet vessels of each model as a proportion of the total flow rate or cardiac output introduced at the aortic root. The MCAs, ACAs, and PCAs, are the middle cerebral arteries, anterior cerebral arteries, and posterior cerebral arteries respectively. The flow rates shown here are representative of sum of the average volumetric flow rate for both the right and left vessels for the ACAs, MCA, PCAs, subclavians, and external carotids.

### 3.5 Circle of Willis Proximal Collateral Flow Routing for Varying Stenosis

In addition to varying extent of flow routing through the cervical vessels feeding the CoW, the communicating arteries in the CoW anastomosis can also recruit flow through them differently based on stenosis severity, so as to retain cerebral blood flow distribution. We investigated this proximal collateral flow routing patterns in the CoW communicating artery segments (*compared to distal collateral circulation through smaller leptomeningeal vessels*), by computing flow rate from simulation data at various locations along the CoW using cross-sectional integration of velocity, and time-averaging over a cardiac cycle (*as described in Section 3.4*). Figure 12 depicts these averaged volumetric flow rates, and the direction of the average flow, through the communicating artery segments in the CoW, for moderate left carotid stenosis paired with moderate right carotid stenosis. Figure 11 depicts the same for moderate right carotid stenosis paired with moderate left carotid stenosis. Additionally, for the 12 CCO cases, the corresponding CoW communicating artery segment flow maps are presented in Figures 13 and 14. Differential flow recruitment and changes in flow routing patterns based on stenosis severity is clearly observed across all of these illustrated flow data. The null hypothesis that communicating artery flows are same as those in the case of flow without any stenosis present was rejected with a p-value *<* 0.01 (*using a one-sample Wilcoxon t-test*). Specifically, for instance, observing the cases with moderate left carotid stenosis paired with severe right carotid stenosis, the anterior communicating artery (AcoA) switches direction, and AcoA flow increases towards the right with increasing right carotid stenosis severity. This trend is conversely true for cases with moderate right carotid stenosis paired with severe left carotid stenosis. Furthermore, for all CCO cases, the AcoA flow stays oriented towards the hemisphere ipsilateral to the occluded carotid. As another instance, we observe that with increasing flow recruitment through the Basilar Artery, the flow routed through the right P1 communicator (RP1) segment increases steadily with extent of right carotid stenosis severity (*and likewise for flow through the left P1 communicator for increasing left carotid stenosis severity*). This is also noted in the CCO cases, where with increasing severity of the left/right carotid stenosis, in presence of fully occluded right/left carotids, flow through the basilar artery into the LP1/RP1 segments increase. Indicating the inherent right-left asymmetry, we also note that flow through the right posterior communicator segment (R.Comm) switches direction for the two most severe right carotid stenosis models to compensate for RMCA and RACA flow while right carotid supply into the CoW is significantly reduced. Together, these results potentially illustrate pathways through the proximal collateral circulation at the CoW that can promote non-intuitive thromboembolus transport respective to the stenosis location and severity.

**Figure 11:**
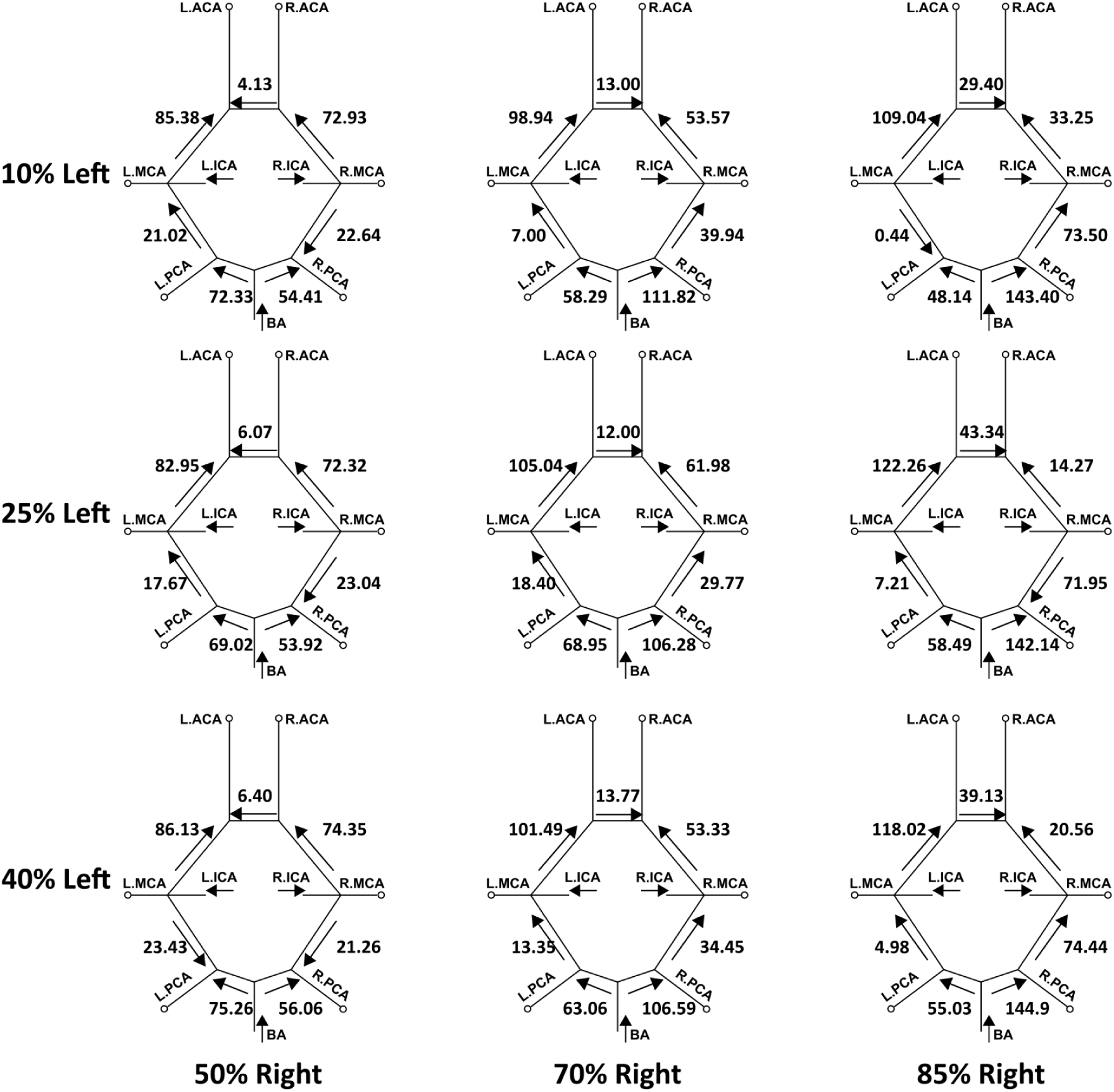
A mapping of the average volumetric flow rate (mL/min) through each communicating artery across all simulated models that have a mild/moderate left carotid stenosis and a severe/high-moderate right carotid stenosis. Each diagram presents the flow rate magnitude and direction for the specific stenosis model pairing, indicating differential flow recruitment and routing of flow across the communicating vessels of the Circle of Willis.

**Figure 12:**
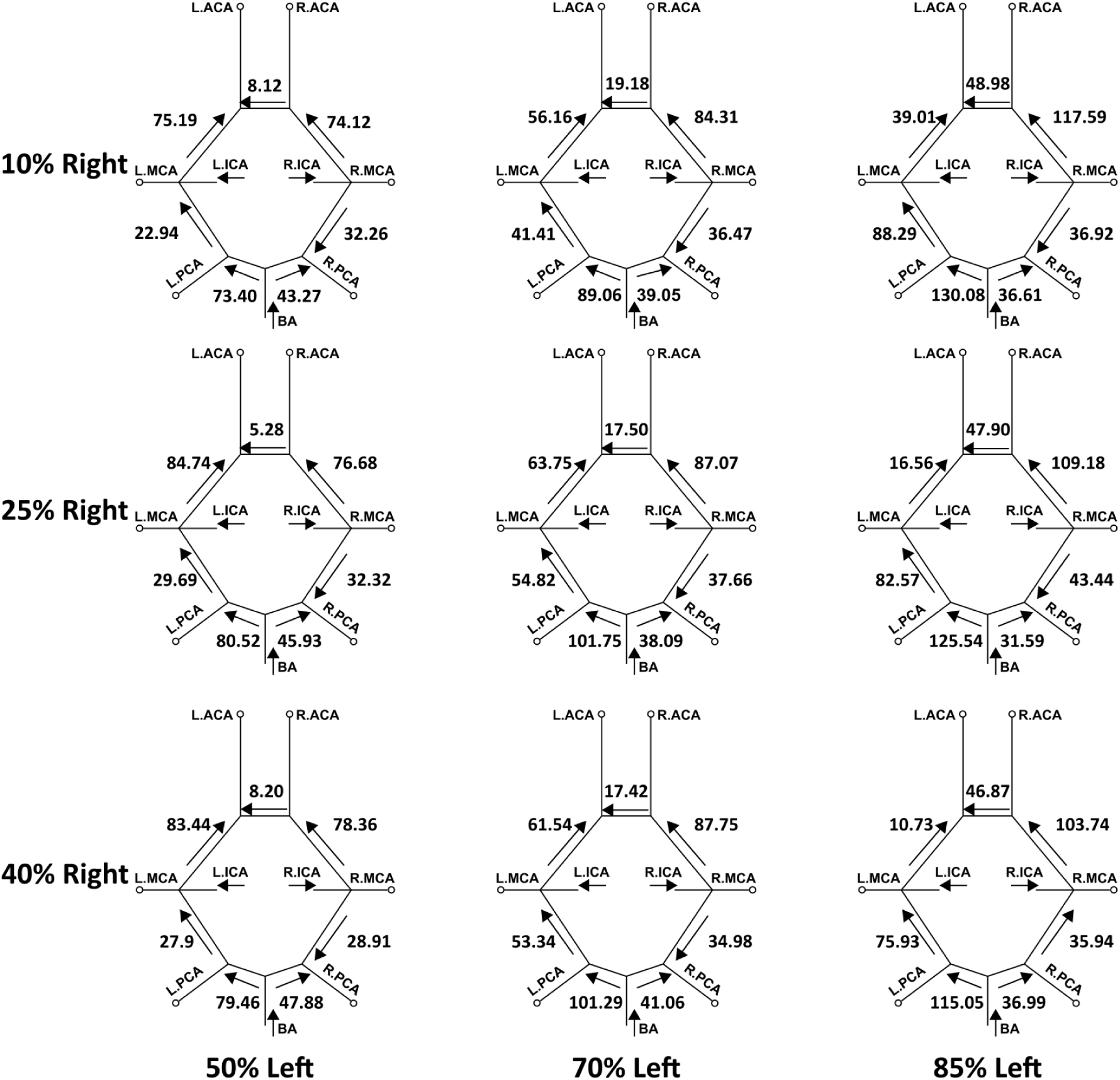
A mapping of the average volumetric flow rate (mL/min) through each communicating artery across all simulated models that have a mild/moderate right carotid stenosis and a severe/high-moderate left carotid stenosis. Each diagram presents the flow rate magnitude and direction for the specific stenosis model pairing, indicating differential flow recruitment and routing of flow across the communicating vessels of the Circle of Willis.

**Figure 13:**
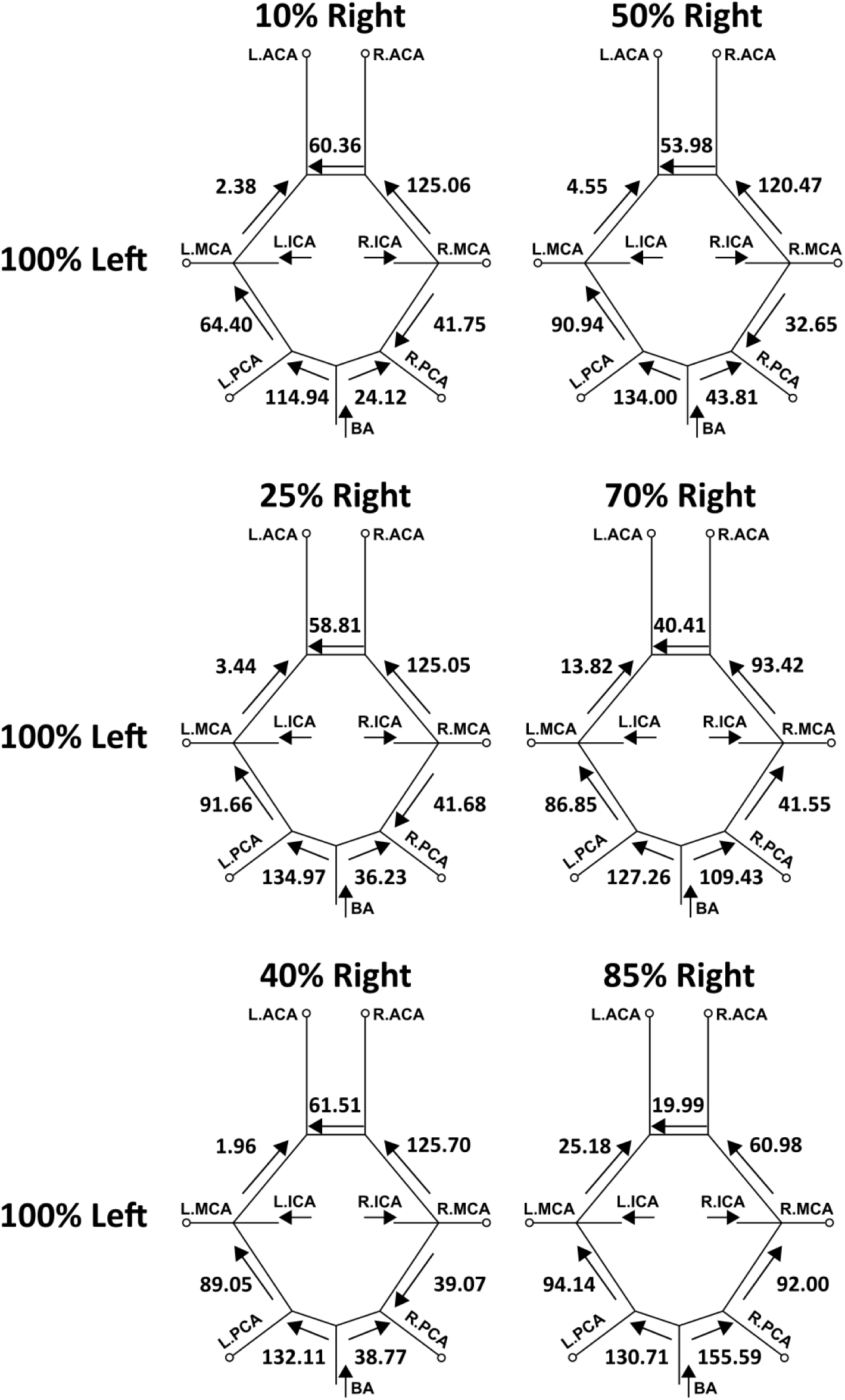
Average volumetric flow rates (mL/min) for each of the communicating segments in the Circle of Willis within the models including a fully-occluded left carotid (CCO) paired with a right carotid with the respective severe and non-severe stenosis degree.

**Figure 14:**
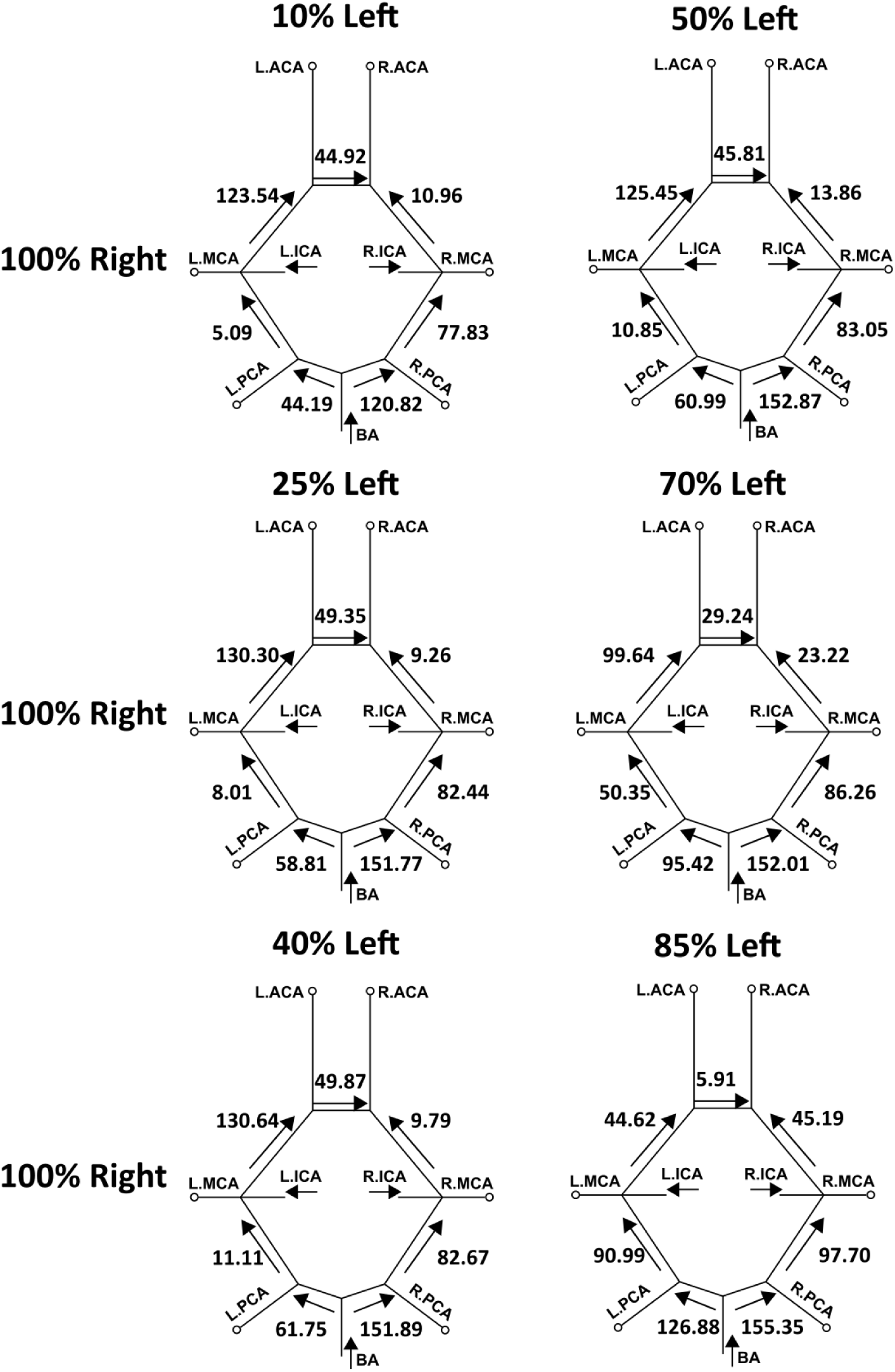
Average volumetric flow rates (mL/min) for each of the communicating segments in the CoW within the models including a fully-occluded right carotid (CCO) paired with a left carotid with the respective severe and non-severe stenosis degree.

## 4 Discussion

### 4.1 Mechanisms of Non-Intuitive Embolus Transport from Carotid Arteries

Our investigation comprised a set of parametric simulations inquiring how thromboembolus transport is influenced by varying combinations of stenosis severities to specifically illuminate how stenosis degree may contribute to elusive or non-intuitive stroke etiologies through the mechanism of contralateral or trans-hemispheric embolus movement. Fundamentally, vascular pathways feeding into the brain comprise a complex network. Flow can be routed through varying, often redundant, pathways to the major cerebral artery territories, in the event of a pathological disruption to flow in one or more vessels in the network. This manifests in differential flow recruitment in the cervical vessels, and proximal flow routing in the communicating vessels of the CoW. While the role of CoW vessels in promoting proximal collateral flow routing for acute ischemic stroke has been discussed in several works [34–37]; the explicit role of CoW vessels in routing emboli in the brain, and driving ambiguity in source-destination relationship has been sparingly investigated. In a series of prior works, we have generated some of the first evidences on how embolus interaction with these varying hemodynamic pathways ultimately determines embolus fate [18, 20]. Here, we further expand upon this observation, and substantiate the role that proximal collateral flow routing may play in underlying non-intuitive embolus distribution in the brain from carotid artery sites.

For trans-hemispheric contralateral embolus distribution, a common pattern seen in our simulation data was that the majority of contralateral embolus movement occurred from the mildly stenosed carotid source to the hemisphere with the respective severe stenosis degree (*two-sample, Mann-Whitney U test between contralateral movement from the severe carotid release and the mild carotid release: p-value <* 0.01). This pattern can be interpreted based on the altering proximal collateral flow routes in the CoW that are dependent on the bilateral pairing of stenosis severities. Based on observations presented in Sections 3.1, 3.3, 3.5, there are three major flow alterations seen for increasing severity of the more severely stenosed carotid (*that is, 10L50R, 10L70R, 10L85R*), where the AcoA and posterior communicators play a key role (*as noted in Section 3.5*). First, the AcoA flow directs from the CoW hemisphere fed by a mild ipsilateral stenosed carotid to the contralateral severe carotid as the severe stenosis increases in degree. The AcoA flow rate also increases as the severity of the moderate/severe carotid stenosis degree increases. These two factors raise the potential of the AcoA recruiting emboli and enabling trans-hemispheric contralateral movement from the relative mild carotid hemisphere to the severe carotid hemisphere due to the favorable AcoA flow rate and flow direction. Second, the flow rates in the internal carotid artery feeding into the CoW increase in the hemisphere on the mild carotid stenosis side, and decrease in the hemisphere on the severe carotid stenosis side, allowing for more emboli to be recruited into the anterior region thus raising the probability of emboli being pulled into the AcoA. Third, as the stenosis severity increases, flow in the P1 communicating artery on the hemisphere ipsilateral to the mildly stenosed site increases and directs towards the anterior region. This is due to flow compensation from the basilar artery for the severely stenosed region. Basilar artery carries flow into the posterior region of the CoW, thereby creating an overall increase of blood flow within the P1 and Comm. arteries in that respective hemisphere.

The CCO cases with fully occluded carotid arteries follow a similar pattern, with observably more pronounced contralateral or trans-hemispheric flow and embolus routing. The AcoA notably plays a role here too in enabling thromboembolus movement from the hemisphere on the partially occluded carotid side to that on the fully occluded carotid side. Similar to non-CCO cases, flow routes through the cervical vessels to the side that has the severe or complete stenosis to compensate for the lack of flow, enabling the propensity of trans-hemispheric movement. Extrapolating purely based on non-CCO cases, we would expect then that complete carotid occlusions would increase the extent of contralateral severity, as more flow enters the CoW from the vertebrobasilar vessels, and the carotid that is not fully stenosed, which is also in accordance with observations presented in Section 3.3. We note that although individual patient vascular anatomical features such as vessel diameters would play a heavy role in the distribution of embolus as also illustrated in our prior works [18, 20], the results here indicate that hemodynamic changes such as differential flow-routing at the network-level induced by the presence of stenosis play an influential role in embolus transport to the cerebral arteries, in addition to individual anatomical variations.

### 4.2 Effects of the Hemodynamic Network Flow Compensation on Cardiogenic Embolus Distribution

We also investigated cardiogenic emboli distributions to the CoW for varying stenosis severity to gain insights on source-destination propensities for cardiac vs carotid sources of thromboemboli. As the severity of carotid stenosis increases on one side (say, left), the less stenosed carotid on the other side (say, right) and the basilar arteries compensate for flow to maintain cerebral perfusion. As stenosis severity on the other side vary, resistance to flow through the stenosis overpowers, shifting the compensation balance further towards basilar artery flows alone. Consequently, again, a network based flow compensation effect, leads to observations on rather consistent cardiogenic embolus distributions across stenosis values in the non-CCO cases at an average of 55% and 45% to the left and right hemispheres respectively for the non-fully-occluded cases. This was also observed for the CCO models whereas there was a consistent distribution for both the left and right occlusion across the varying severities as seen in Figure 7.

These observations on contralateral thromboembolus distribution are mechanistically informed by the mildly stenosed carotid and the basilar artery recruiting more flow, and more emboli, towards the CoW, and subsequent trans-hemispheric transport of these emboli based on CoW proximal routing noted above pushing a fraction of these emboli into the hemisphere associated with the more severely stenosed carotid. Figure 15 presents a conceptual schematic summarizing these two effects, discussed extensively thus far, originating from two specific flow recruitment patterns in the cerebrovascular network: differential recruitment of flow at the cervical vessels, and proximal collateral flow routing at the CoW. This summary figure schematically illustrates a case of severe right carotid stenosis experiencing contralateral movement of carotid emboli, and non-intuitive transport of cardiogenic emboli, to the right hemisphere Supplementary animation V6 provides a visual representation of these effects occurring in the CoW. From this perspective, for cardiogenic emboli with varying degrees of stenosis, we observe limited distinct preference in laterality of distribution for the simulations considered. Consequently, for a stroke patient with indicated carotid disease and suspected cardioembolic source, these results indicate that in the absence of quantitative source-destination transport patterns, disambiguating between cardiogenic and carotid etiologies would be challenging, which is commonly observed in ESUS cases.

**Figure 15:**
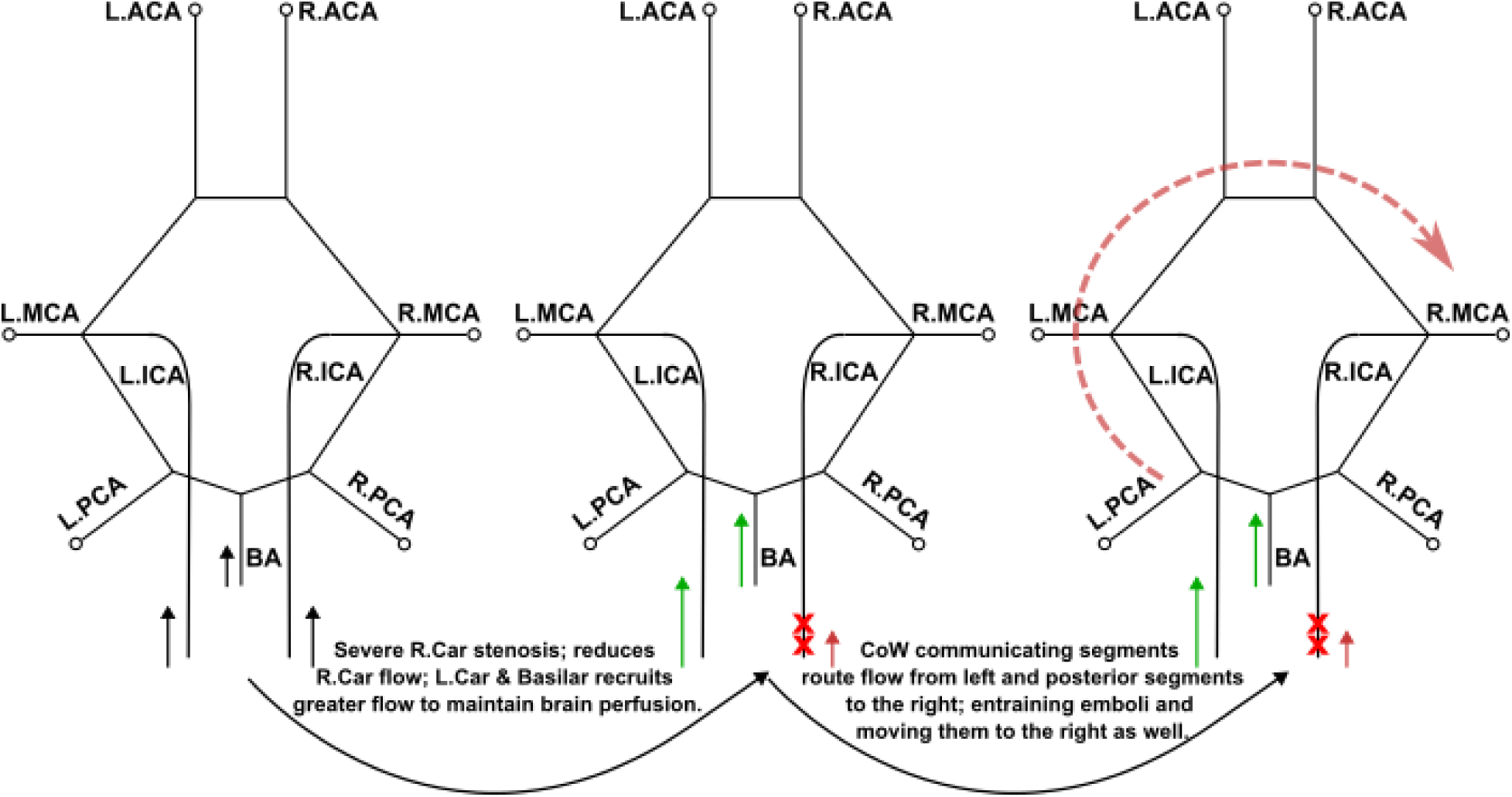
A schematic concept sketch that aims to summarize the observations on contralateral embolus source-to-destination transport based on potential hemodynamic interactions due to differential flow recruitment through the basilar artery and less stenosed carotid artery, and due to proximal collateral flow routing at the Circle of Willis

Our results specifically illustrate how the proximal collateral flow of the CoW influences distribution of embolus to the cerebral arteries. These additional results presented here help to illuminate how emboli are transported into both of the common carotid arteries but do not directly reflect the distribution of embolus into either hemisphere of the CoW. It is commonly understood that the right common carotid artery has a higher susceptibility of recruiting cardiogenic embolus that is reflected in our results shown here in Figure 5. Although there is a preferential transport to the right common carotid, this is not reflected at the level of cerebral distribution as there is a 55% and 45% average of cardiogenic embolus to the left and right CoW hemisphere respectively for the partially-occluded models (Figure 4). This helps to highlight the importance of the proximal collateral flow and the basilar flow compensation in the presence of varying stenosis as the laterality of embolus uptake at the common carotid arteries does not fully dictate the hemisphere distribution at the cerebral arteries. Additionally, Figure 5 acts a validation effort of our embolic transport framework in that cardioembolus have a preferential pathway into the right common carotid as observed in prior clinical studies [32].

### 4.3 Clinical Relevance

The findings from this study provide critical insights on embolus-hemodynamics interactions, and embolus source-destination relationship, in the context of deciphering etiology and intervention targets in potential embolic events with carotid stenosis. As per current standard-of-care, carotid intervention is indicated when stenosis severity is greater than 70% in the ipsilateral carotid along with the correlating symptoms (*symptomatic*) [7]. Yet, there is substantial evidence that stenosis severity is not the only determinant for stroke risks, and less severe stenosis sites can lead to stroke or embolic events [38, 39]. Findings from our study indicate potential mechanisms driving an increased risk of embolus transport from a mildly stenosed contralateral carotid in the presence of severe ipsilateral carotid disease. This provides the basis for scenarios where carotid interventions could be indicated for the contralateral carotid to prevent stroke recurrence. Furthermore, in the presence of an ipsilateral carotid occlusion (*both partial and complete*), we show small but significant levels of trans-hemispheric embolus transport from the contralateral carotid. This finding could support consideration of intervention on the contralateral carotid for secondary stroke prevention in patients with chronic carotid occlusions. Our completed simulations show that up to 14% of embolization from the carotids can lead to contralateral embolic events that may keep a carotid that is symptomatic or actively embolizing from being properly identified. Through our findings, non-intuitive trans-hemispheric contralateral embolus distrbution is shown to have the potential to pose a prominent risk, and further risk stratification may be beneficial in deducing treatment for patients that are experiencing symptomatic carotid stenosis with a corresponding stroke. Furthermore, often carotid stenosis may co-exist with cardiac factors leading to embolisms, causing the co-occurrence of more than one likely sources which makes disambiguating stroke etiology a challenge. This is the hallmark of ESUS cases, and findings form our study shows that detailed understanding of embolus source-to-destination distribution can help address this underlying challenge, explaining non-intuitive embolus distribution pathways. Understanding how a stenosis site can contribute to an embolic event in the brain, especially in presence of another major co-occurring embolsim source like the heart, is therefore of high clinical importance. Yet such quantitative information on embolus source-to-destination transport remains currently impossible to obtain using standard-of-care imaging techniques or animal models, thereby making the computational findings reported here notably significant.

## 4.4 Assumptions and Limitations

This study was based on a few key assumptions. The underlying computational model relies on several assumptions to make high resolution simulations such as the ones presented here tractable, which have been discussed at length in our prior works [18, 20]. We assume that blood flow influences the emboli, but the emboli themselves do not significantly alter the flow. Each embolus is modeled independently, and does not interact with other emboli, and embolus sizes are small relative to the size of the arteries modeled; making it reasonable to assume one-way interaction of blood flow and embolus. For heart-to-brain cerebrovascular models, this assumption can be further improved upon, when considering larger emboli traversing through the cerebral arteries, where the differences between embolus size and vessel diameter may be smaller. Additionally, small embolus sizes avoid numerical issues with clogging the stenosis region, as we assumed a rigid particle and squeezing deformations are not possible within our simulation scope. Second, we considered only a single embolus size in our study. This a meaningful simplification because we have extensively demonstrated how size-dependent embolus-hemodynamics interactions manifests in embolus source-to-destination distribution in a number of prior works [18, 20, 23, 27]. Hence, controlling the simulations to one embolus size enabled us to focus the computational efforts on stenosis severity and network flow routing effects which have received less attention in prior efforts. Our goal also was to unravel source to destination movement of emboli, and not to incorporate the propensity of embolus to form at a stenosis site. While stenosis severity and plaque size is one factor determining embolism risks, the fundamental variable governing embolic events is plaque vulnerability. Identifying a vulnerable plaque comes with some nuance where some larger plaques can be calcified and have less of an embolization threat than one that is more of a mild-moderate stenosis degree [40, 41]. Developing plaque rupture and embolization models, and combining them with a embolus transport study as presented here, is a substantially expensive computational effort that was beyond the scope of this study. Furthermore, this focused analysis enables us to understand the source-to-destination transport patterns for small emboli which plays a critical role in a range of complications such as microstrokes, lacunar and watershed infarcts, and cerebral small vessel disease, for which embolus etiology is often harder to disambiguate. Likewise, we used one base patient anatomy that was modified across multiple stenosis severities which allows for controlled anatomy within the simulations. Here, we did not consider anatomical variations across multiple patient vasculature models. Our focus here was on the hemodynamics mechanisms that underlie non-intuitive embolus distribution, such as flow compensation through less stenosed arteries and proximal collateral flow redistribution in the presence of carotid stenosis. These mechanisms would be at play irrespective of specific patient anatomical variations. A direct extension of this *in silico* analysis developed here would be to directly recreate stenosis geometries and inform potential release sites for a larger cohort of patients from images, and estimate thromboembolus distribution probabilities. Third, we have not accounted for specific cerebrovascular circulation features such as cerebral autoregulation and distal collateral flow for our study. This is primarily to capture general parametric trends with stenosis severity while maintaining the overall computational costs low. Integration of robust yet computationally efficient models for such processes within the *in silico* framework presented here remains an active area of investigation. Additionally, changes in distal vascular resistance values due to the presence of stenosis were not accounted for within our framework as we wanted to consider the hemodynamic changes within the network assuming regular perfusion to the arterial beds. Furthermore, empirical resistance values for our parametric framework are not readily available without clinical or benchtop studies that can integrated with *in silico* experimentation, which is a substantially complex endeavor. Fourth, we considered the stenosis region to occlude the same percentage of cross-sectional diameter and no remodeling to occur after embolization. We are less concerned with embolization and more so embolic transport where we assume the stenosis region is displaying levels of continuous shedding of smaller particle that can cause downstream, cerebral occlusions versus full breakage in which occlusion degree remains constant. Finally, our model focused on distribution trends for thromboemboli only. While stenosis sites can cause thrombogenesis and subsequent thromboembolization, ruptured plaques can often release cholesterol atheroemboli. Our analysis presented here can be directly extended to study cholesterol atheroemboli and atheroembolic showers in future investigations.

## 5 Concluding remarks

We have presented an *in silico* investigation on how varying degrees of bilateral carotid stenosis can induce non-intuitive embolus transport within the cerebral arteries that can ultimately pose challenges with disambiguating etiology and confusing treatment or intervention decisions. A total of 78 simulations were completed with 18 models of varying bilateral moderate/severe stenosis degrees and 12 models with contralateral carotid occlusion cases (CCO) with pairing of differing stenosed carotids on the non-occluded side. Contralateral embolus movement is the primary mechanism of non-intuitive transport within our simulations and all cases experienced a non-zero contralateral source-to-destination transport of thromboemboli. Models paired with the most severe partial occlusion degree (85%) were found to manifest the highest likelihood of contralateral thromboembolus movement, with fully-occluded CCO cases having on average a higher probability of contralateral movement than non-CCO cases. Cardiogenic thromboemboli did not exhibit a distinct hemisphere preference among all models, with there being consistent distributions to each side of the models regardless of stenosis degree due to flow compensation of the basilar artery and the less stenosed carotid artery. This was consistent for both non-CCO and CCO cases. Ultimately, these findings illustrate the capability of non-intuitive embolus travel from cardiogenic and contralateral carotid sources such that carotid embolus can seemingly source from the opposite side of severely stenosed carotid site, while cardiogenic emoblus can move to either carotid artery. We demonstrate the potential underlying hemodynamics based mechanism for this contralateral movement, comprising differential flow routing in the cervical vessels feeding the Circle of Willis, and proximal collateral flow routing across the communicating artery segments of the Circle of Willis. With procedures such as CEA having a dependence on occlusion location, these findings can be beneficial for optimizing both treatment efficiency and efficacy in reducing recurrent events.

## Supporting information

V1-StenosisLocalFlow

V2-40R85L-AllEmbolusReleases

V3-ContralateralMovement

V4-ContralateralMovement-DualView

V5-CompleteOcclusionTransport_DualView

V6-CoWFlow

## Conflicts of Interest

The Authors RR, ML, NJ, and DM have no conflicts of interest to report regarding this study.

## Funding Acknowledgements

The Authors acknowledge funding support from National Institutes of Health Award: R21EB029736 for this study. This work utilized the Summit supercomputer, which is supported by the National Science Foundation (awards ACI-1532235 and ACI-1532236), the University of Colorado Boulder, and Colorado State University. The Summit supercomputer is a joint effort of the University of Colorado Boulder and Colorado State University. The funders had no role in study design, data collection and analysis, decision to publish, or preparation of the manuscript.

## Author Acknowledgements

RR, and DM designed the study; RR conducted all simulations and data analysis; DM, NJ, and ML interpreted simulation data; NJ and ML provided clinical insights; RR drafted the manuscript along with DM; RR, NJ, ML, and DM reviewed and edited the manuscript. All authors agreed to final version of the manuscript.

## Data Availability

All quantitative embolus distribution data for this study have been shared through the Open Science Framework as part of project titled, “Dataset: Transport and Distribution of Embolic Particles in Human Vasculature,” which can be accessed at: https://doi.org/10.17605/OSF.IO/CQKZT. All image-based modeling and computational hemodynamics simulations were conducted using the open source software package Sim-Vascular. The image-based models (segmentations, pathlines) and computational hemodynamics data will be made available through the Vascular Model Repository associated with the SimVascular project and can be accessed at https://simvascular.github.io/. The sharing process with Vascular Model Repository is ongoing, and when the models have been shared, they will be linked to the dataset released on Open Science Framework linked above. FlowVC was utilized for completing embolic simulations where the source code can be found at: https://shaddenlab.berkeley.edu/software.html. Additionally, statistical tests utilized in this experimentation has been provided to PLOS Computational Biology. For any data access issues, or for additional access to scripts and tools, please contact the corresponding author directly via email, or via the contact page on the research team’s official website: https://www.flowphysicslab.com/.

## Supplementary Material Information

## S1 Animation of stenosis flow and embolus dynamics

Here, we describe the simulation animations that have been provided as additional illustration of the simulation data to support discussions presented in the main manuscript.

Animation titled: ***V1-StenosisLocalFlow.mp4*** shows the flow disturbance caused by the severe stenosis degree of 85% across all three moderate stenosis degrees. This flow disturbance results in a jet impingement that varies in length with respect to the contralateral carotid stenosis degree. This stenosis results in differing carotid embolus recruitment from the cardiogenic source and altered downstream embolus dynamics.

Animation titled: ***V2-40R85L-AllEmbolusReleases.mp4*** shows how all three embolus releases travel into the cerebral arteries from respective surface, presented for the non-CCO 40% stenosed right carotid with 85% stenosed left carotid case. Dynamic source-to-destination transport patterns, trends in embolus recruitment in the vertebrobasilar pathway, and contralateral movement into the anterior CoW vessels can bee seen in the video.

Animation titled: ***V3-ContralateralMovement.mp4*** shows one instance of how contralateral trans-hemispheric embolus movement can be enabled through the AcoA, with emboli moving from the right carotid release to the left hemisphere. This for the case of 10% right carotid stenosis paired with 85% left carotid stenosis.

Animation titled: ***V4-ContralateralMovement-DualView.mp4*** shows thromboemboli released from two sources, both from the right carotid but of differing stenosis severity. This animation illustrates how the likelihood of contralateral movement is higher in cases with higher severity pairing where the amount of particles traveling into the left hemipshere is visibly higher in the model with a 40% right severity and a 85% left severity.

Animation titled: ***V5-CompleteOcclusionTransport-DualView.mp4*** compared two differing cases of CCO with one having a 10% severity and the other 85%. Both the cardiogenic and carotid releases are shown where we can note two distinct mechanics: (1) the recruitment of cardiogenic embolus through the vertebral arteries that supplements the occluded and highly stenosed carotid arteries, (2) the distinctly different hemodynamics within the 85% stenosed carotid where the embolus movement is much more erratic and less intuitive due to lack of flow and the jet impingement.

Animation titled: ***V6-CoWFlow.mp4*** compares two stenosis models where there is a mild 10% stenosis degree in the left carotid and a moderate degree of 50% in the right carotid presented in the left column and a severe stenosis degree of 85% in the right carotid presented in the right column. This animation compares flow rate directioj and magnitude within the communicating arteries that make up the collateral cerebral cirulation. We can see distinct differences between these cases in: (1) higher flow rates within the posterior and anterior region for the severe case (specifically the left internal carotid extension member, the right p1 connector, and the AcoA), (2) flow direction within the same vessels re-route towards the severe-hemisphere from the 10L50R case to the 10L85R with a decrease in the right internal carotid extension flow rate. These characteristics of collateral flow routing and magnitude changes allow for increased contralateral movement and non-intuitive movement within certain cases. This animation acts supplements Figure 9 showing the flow pathways that allow for embolus routing from the hemisphere ipsilateral to the mild/moderate stenosis to the contralateral, severe carotid stenosis hemisphere.

## S2 Mathematical model details for hemodynamics simulation

Here we present the foundational equations for the stabilized finite element hemodynamic simulations. Blood was modeled as a Newtonian fluid with a bulk density of 1.06 g/cc and viscosity of 4.0 cP. whose momentum and mass balance obeys the standard Navier-Stokes and continuity equations stated in Eq. S1 and S2 as stated below:

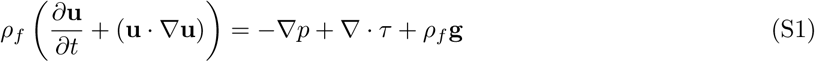

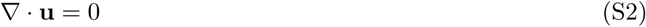

The term *τ* represents the viscous stresses in blood, which is modeled as per a linear Newtonian stress vs strain rate relation as follows:

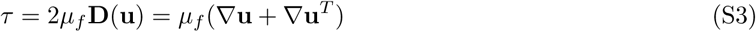

These equations are solved using matrix systems of equations obtained by decomposing a stabilized Petrov Galerkin finite element variational formulation of these equations over a mesh composed of linear tetrahedral elements. This variational formulation is stated in Eq. S4 as follows:

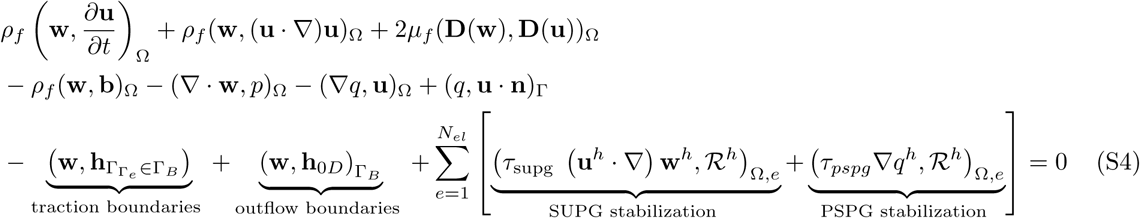

In these equations, **u**, p are blood flow velocity and pressure, **w**, q are the two test functions employed in the finite element solver, *µ_f_* is the averaged blood viscosity, *ρ_f_*is the averaged density of blood, **g** designates gravity, Ω denotes the computational domain of the arterial network, Γ and the subscripts of Γ denote the boundary faces of the computational domain, R*^h^* is the residual of the momentum equation, and *τ_supg/pspg_* are the SUPG stabilization and PSPG stabilization factors used for convection and pressure stability respectively. **h**_0*D*_ represents the terms used to integrate the boundary conditions at the outlets and inlets of the arterial network; while 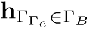 represents the other boundary conditions that may be necessary to run physiologically realistic flow simulations.

## S3 Mathematical model details for embolus dynamics simulation

Embolic particle simulations were conducted using a custom-modified version of the Maxey-Riley equation stated in Eq.S5 as below:

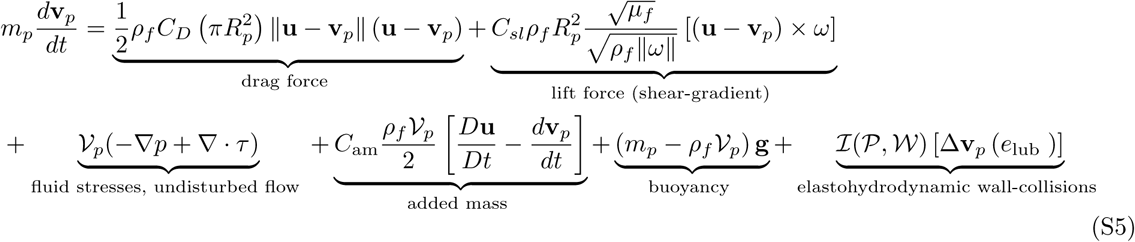

where m*_p_* notes the particle mass, V*_p_* is the particle volume, R*_p_* is the particle radius, and **v***_p_* is the translational particle velocity. C*_a_m* is the added mass coefficient, C*_D_* is the drag coefficient, and C*_s_l* denotes the shear-gradient lift force. *ω* = ∇x **u** is the vorticity of the flow where the particle is located. *I*(*P*,*W*) is an indicator that has a value of 1 when a particle *P* comes into contact with the artery wall, the value of this function is 0 otherwise. Next, the contribution of the particle-wall collision to the change in particle velocity is denoted by the term [Δ**v***_p_*(e*_lub_*)], which accounts for elastohydrodynamic lubrication effects at the collision point. The term *e_lub_* is the restitution coefficient. Figure S1 illustrates the seeding locations of embolus across the carotid and cardioembolic releases and the offset distance between the arterial wall and embolus that is equivalent to the radius of the embolus.

**Figure S1:**
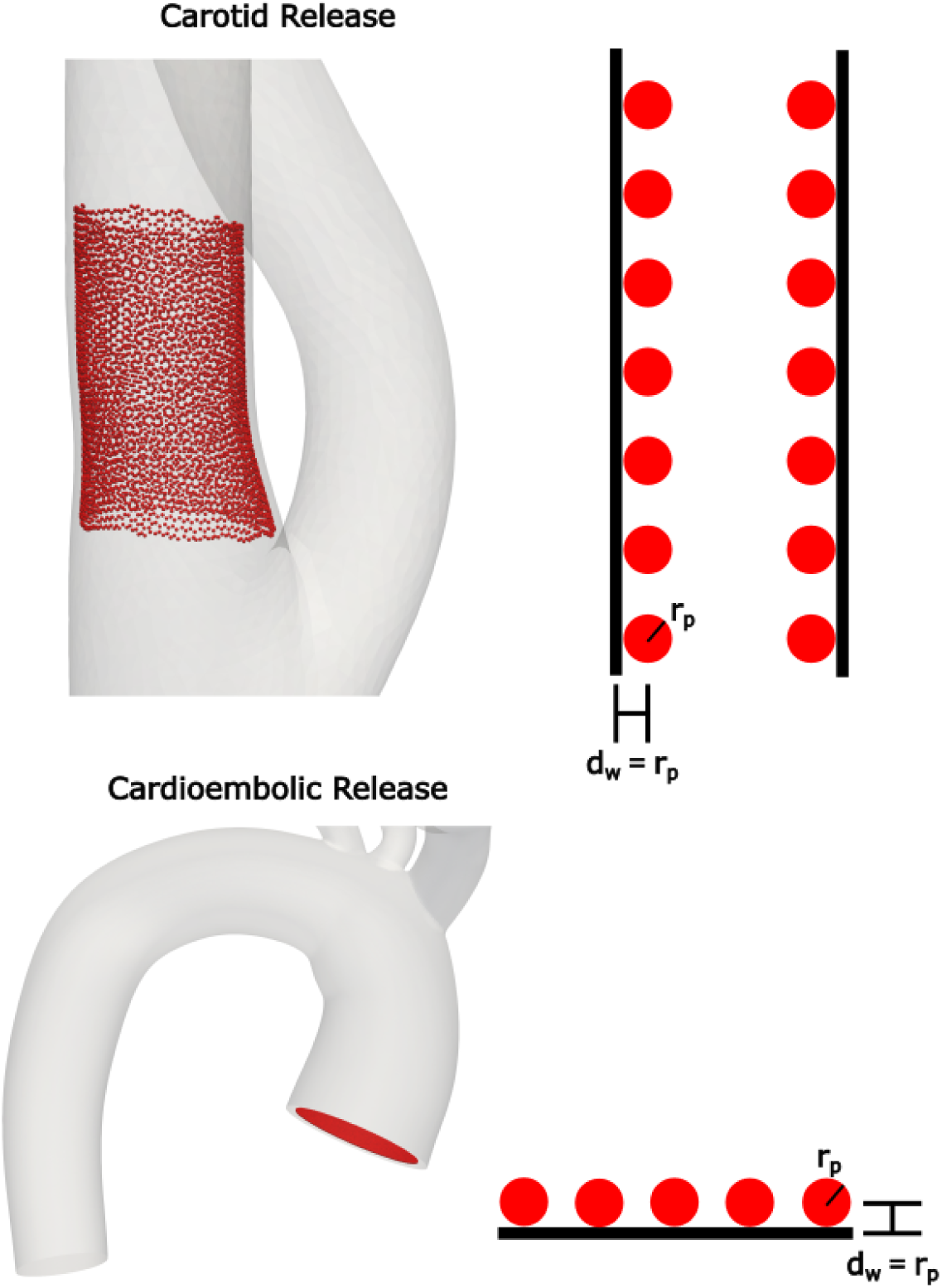
Illustrates how particles are seeded at both the cardioembolic and carotid sources. Embolus are offset from the wall by factor d_w_ that is equal to the radius of embolus considered. For all cases considered in this study, a particle size of 500 microns was used with a d_w_ of 250 microns across all releases.

## S4 Length of stenosis region

The length of the stenosed region across the span of the carotid artery was calculated using the equation below:

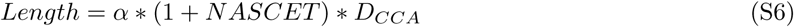

where *α* is a constant factor used across all stenosis applications as described in [42]. NASCET is the associated NASCET stenosis degree applied for that model. *D_CCA_* is the diameter of the undisturbed common carotid artery at that ipsilateral carotid artery. This relation, for a chosen NASCET severity, was used to reduce the segmentation diameters and re-loft the models to create stenosed carotid artery models.

## S5 Complete distribution dataset for emboli across all vessels

For each simulation, the total proportion of embolus samples released was calculated across each outlet of our heart-to-brain model. All embolus source-to-distribution statistics reported in the manuscript were derived from these distributions. While in the manuscript the focus was on contralateral trans-hemispheric distribution, the complete set of embolus distribution across all vessel outlets individually is not included in the main manuscript. We provide a visual depiction of this complete dataset for each embolus release source as additional reference in the supplementary figures below. The vessel outlets included the anterior cerebral arteries (ACAs), the posterior cerebral arteries (PCAs), the middle cerebral arteries (MCAs), the subclavians, the external carotids, and the descending aorta (DA).

**Figure S2:**
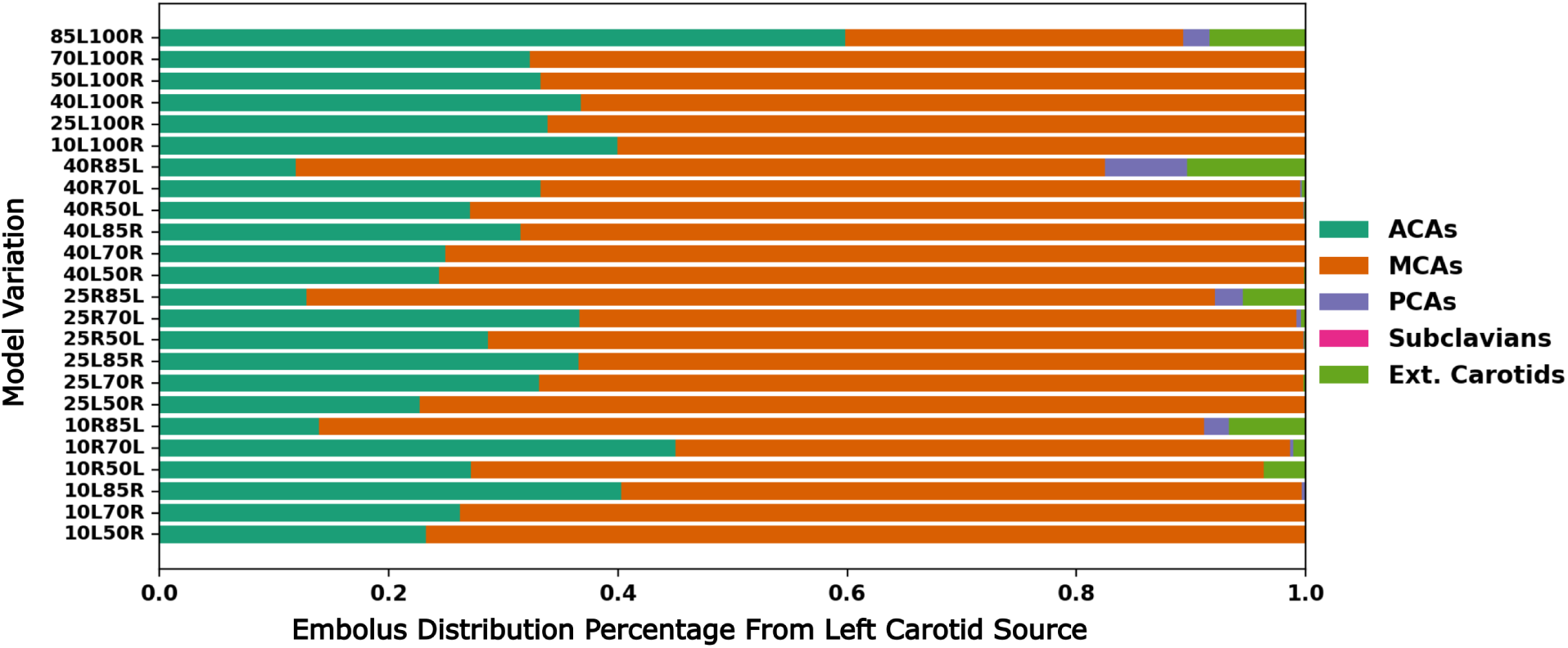
Complete simulated embolus distribution data across all left carotid embolus releases, for all possible vessel outlets in the heart-to-brain pathway modeled for this study.

**Figure S3:**
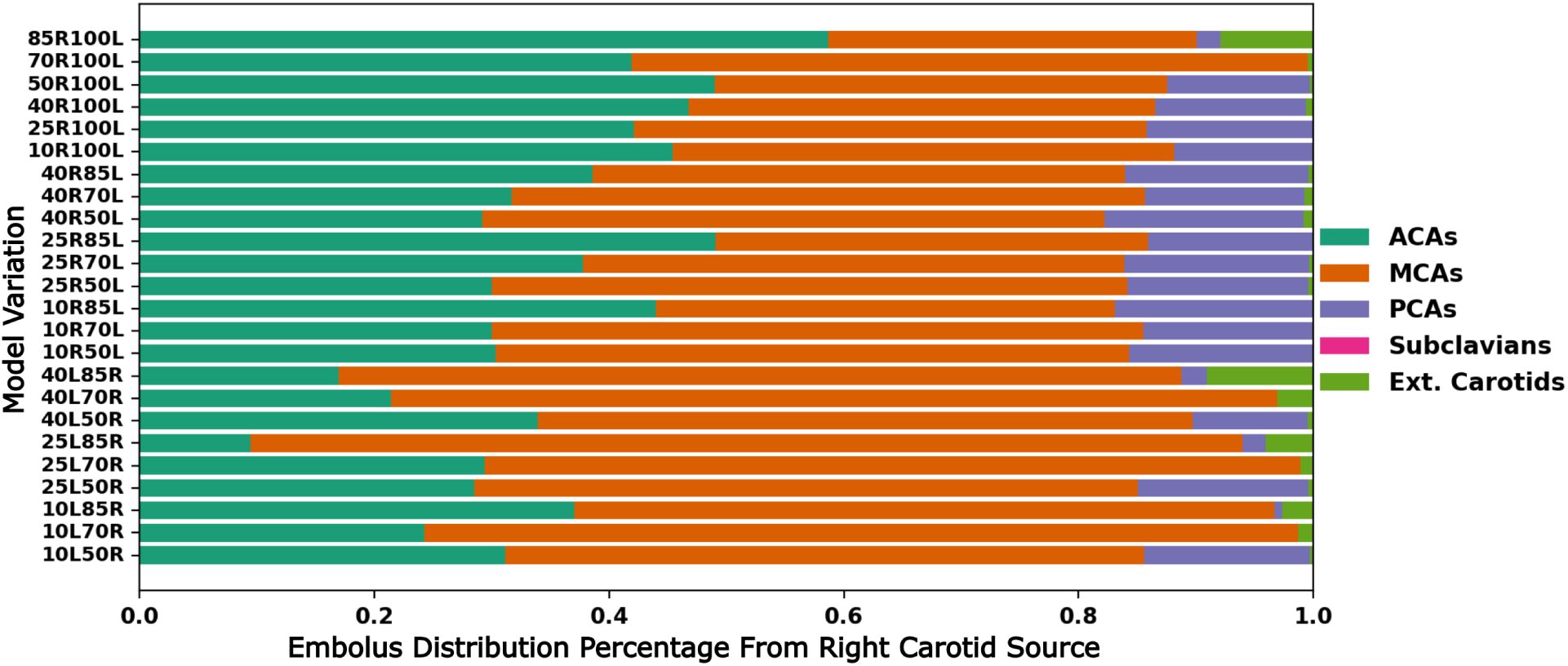
Complete simulated embolus distribution data across all right carotid embolus releases, for all possible vessel outlets in the heart-to-brain pathway modeled for this study.

**Figure S4:**
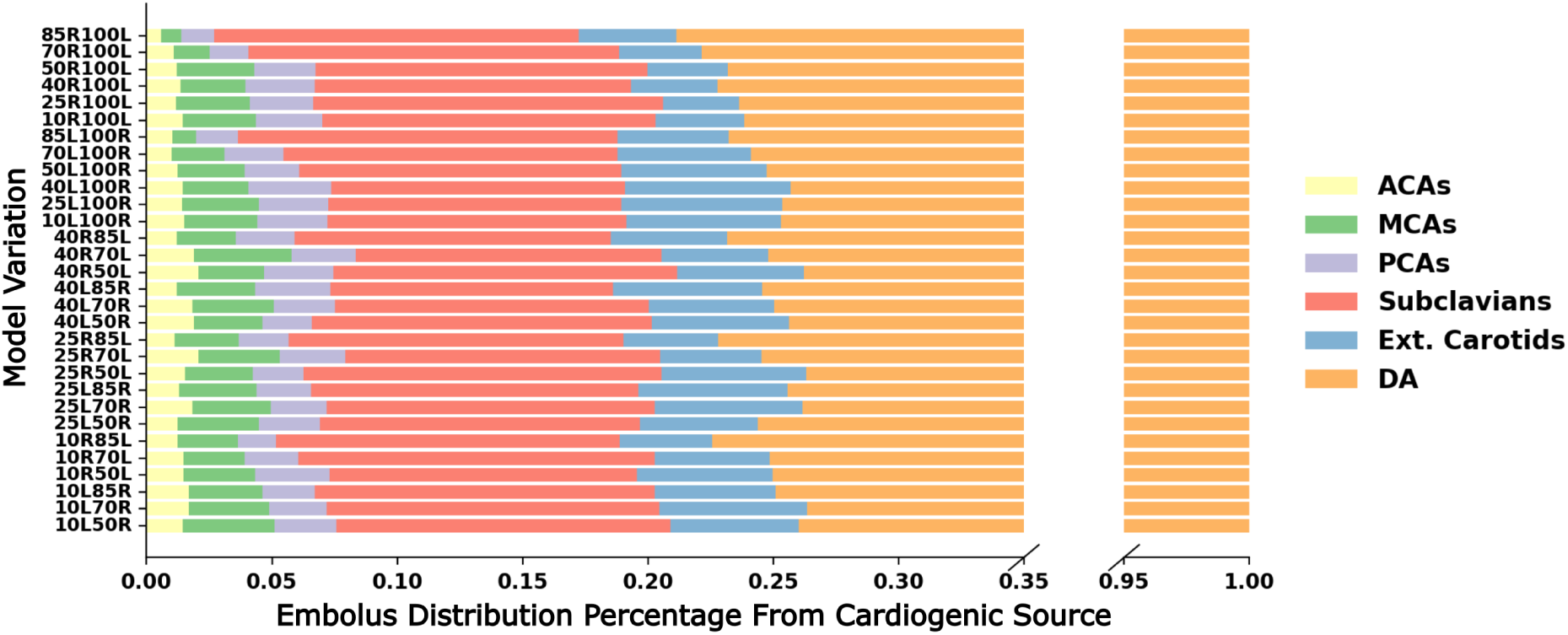
Complete simulated embolus distribution data across all cardiogenic embolus releases, for all possible vessel outlets in the heart-to-brain pathway modeled for this study.

## Notes

### Competing Interest Statement

The authors have declared no competing interest.

### Summary of Updates

We have made significant edits and additions to describe the methodology of the study, including statistical and data analysis aspects. We have also moved several supplementary data and figures into the main manuscript to support and substantiate our arguments on network level flow routing effects.

